# Joint Effects of Early Pandemic Containment Policies on Anxiety in the United States

**DOI:** 10.64898/2026.06.29.26356856

**Authors:** Devon Watts, Pratik Nitin Khadse, Omid Ebrahimi, Justin Tubbs, James Lian, Lorenza Dall’Aglio, Daniel Fatori, Yu Zhou, Pedro Zuccolo, Mihael Cudic, Juan Fernando De La Hoz Gomez, Younga Heather Lee, Gisele Gus Manfro, Sarah Bauermeister, Andre Brunoni, Karmel W. Choi, Chris J. Kennedy, Jordan W. Smoller

## Abstract

**Importance:** The impact of COVID-19 containment policies (e.g., physical distancing, school closures) on population anxiety has been debated and difficult to resolve.

**Objective:** To estimate the joint effects of state-level COVID-19 containment policies on anxiety symptoms during the early pandemic.

**Design:** Retrospective analysis of a prospective cohort with cross-sectional outcome assessment.

**Setting:** *All of Us* Research Program, a U.S. national research cohort.

**Participants:** 40,610 adult participants who completed the *All of Us* COPE survey in July 2020.

**Exposures:** Seven state-level COVID-19 containment policies (school closures, workplace closures, cancellation of public events, restrictions on gatherings, public transport closures, stay-at-home requirements, and restrictions on internal movement) measured from March 22 to May 23, 2020, via the Oxford COVID-19 Government Response Tracker (OxCGRT).

**Main outcomes and measures:** The primary outcome was anxiety symptoms (GAD-7) in July 2020. Using quantile g-computation, we classified policies as anxiety-increasing or anxiety-decreasing by the sign of their training-set contributions, then re-estimated joint effects in a holdout testing set.

**Results:** Among participants (64% female; mean age: 57.8 years), 13.3% (n=5398) reported moderate-to-severe anxiety (GAD-7 score 10-21) in July 2020. The joint effect of all seven containment policies was not significant (β = 1.88, 95% CI: -0.51 to 4.28, p = 0.12). An anxiety-increasing joint effect from 4 policies (school, workplace, public events, internal movement; β = 2.98, 95% CI: 0.30 to 5.66, p = 0.03) and an anxiety-decreasing joint effect from 3 policies (gatherings, public transport, stay-at-home; β = -1.10, 95% CI: -1.75 to -0.44, p = 0.002) reached significance. Effects were largest in adults 18-44 (anxiety-increasing β = 8.93, 95% CI: 1.50 to 16.37, p = 0.02; anxiety-decreasing β = -2.81, 95% CI: -4.98 to -0.64, p = 0.01), with no significant effects in adults 45 and older.

**Conclusions and Relevance:** Modeling seven containment policies jointly showed no net anxiety effect, a result that masked opposing-direction effects. Partitioning by effect direction revealed significant joint effects exceeding single-policy estimates, with young-adult point estimates above the 4-point GAD-7 minimal clinically important difference (MCID) though lower CI bounds fell below it. These findings may inform the use of containment policies in future pandemics, given their differing association with population anxiety.

## Introduction

The COVID-19 pandemic was associated with significant increases in anxiety across multiple countries, with a large global meta-analysis estimating a 25.6% increase in anxiety disorder prevalence during the first year of the pandemic^1^ whereas another reported elevated anxiety symptoms on the Generalized Anxiety Disorder 7-item scale (GAD-7) in 32.06% of the general population^2^. While government containment measures were introduced to curb transmission, their psychological sequelae remain incompletely understood. The risk/benefit trade-off incurred by lockdowns in terms of reducing the spread of disease at the expense of collateral impacts on mental health has been the subject of controversy in the public sphere^3–4^ and the medical literature^5^ and has implications for policy-setting in future pandemics. Stricter containment policies, as captured by the Oxford COVID-19 Government Response Tracker (OxCGRT)^6^ Stringency Index, have been associated with increased psychological distress in a longitudinal analysis of 15 countries.^7^

The relationship between containment policies and mental health outcomes is complex and likely moderated by demographic factors and pre-existing mental health vulnerabilities. For example, younger individuals (<30 years) and those with previously diagnosed mental illness experienced higher levels of psychological distress across countries with varying containment policy responses.^8^ Individual containment policies also appear to show differential impacts on mental health. While higher stringency of physical distancing policies during the early pandemic period was overall associated with worsened mental health, individual policy components revealed heterogeneous effects. Workplace closures, public transport closures, restrictions on gatherings, and stay-at-home requirements were each significantly associated with worsening mental health, while school closures and cancellation of public events showed non-significant associations.^9^ However, these studies examined each policy separately. Since policies were typically enacted concurrently, these individual-policy estimates may not reflect the mental health effects of the policy environment that individuals actually experienced. Prior investigations that assume an equal weighting across policies can also bias effect estimates when policy combinations produce synergistic or antagonistic effects. For example, school closures concurrent with fully operational public transportation may produce social isolation effects different from those observed when examining either policy independently. Joint modeling addresses this by estimating how policies contribute to anxiety while accounting for their co-occurrence.

To address both limitations, we employed quantile g-computation,^10^ an exposure mixture approach that can estimate the joint effect of simultaneously increasing all policy exposures by one quantile (i.e., shifting each policy from its current quartile to the next). Unlike composite indices that assume equal weighting, this method derives empirical weights from the data based on each policy’s association with the outcome. Using this framework, we evaluated seven state-level COVID-19 containment policies on subsequent anxiety symptoms among 40,610 participants from the *All of Us* Research Program^11^ who completed the COVID-19 Participant Experience (COPE) survey in July 2020. Given prior evidence of age-differential vulnerability to both pandemic stressors and policy effects, we also stratified analyses by age group (18-44, 45-64, and ≥65 years).

## Methods

### Study Population

We conducted a retrospective cohort analysis in the *All of Us* Research Program, a longitudinal cohort study that has enrolled participants across the United States (PMID: 31412182). Our source population comprised 254,700 participants with available electronic health record (EHR) data who completed the *All of Us* Basics survey. Our analytic sample comprised 40,610 participants residing across all 50 U.S. states who completed the July 2020 release of the COVID-19 Participant Experience (COPE) survey^12^ and provided valid GAD-7 anxiety scores ranging from 0-21 (Table 1). The survey, with anxiety measures, was administered monthly in May, June, and July 2020.

**Table 1.**
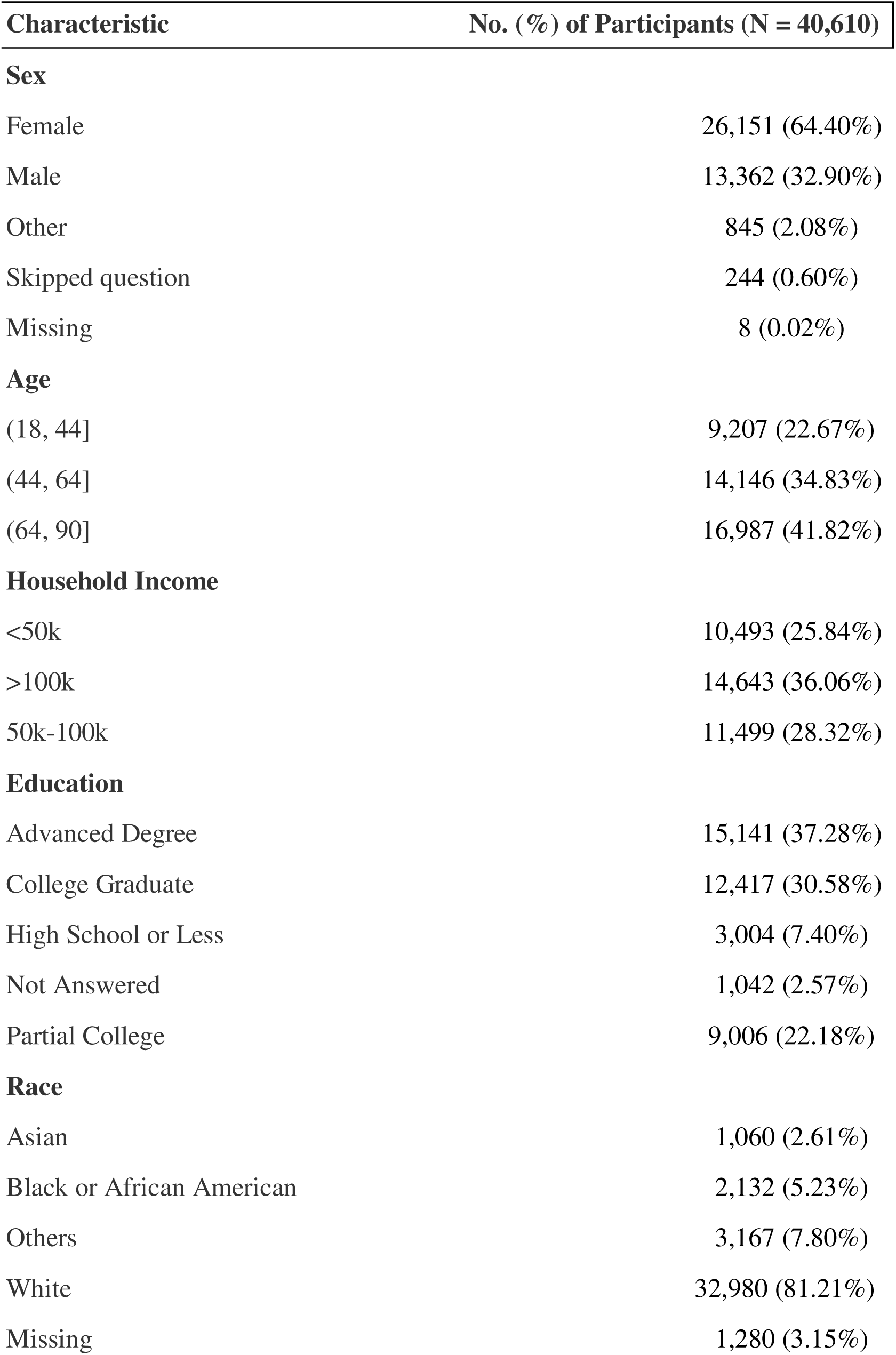
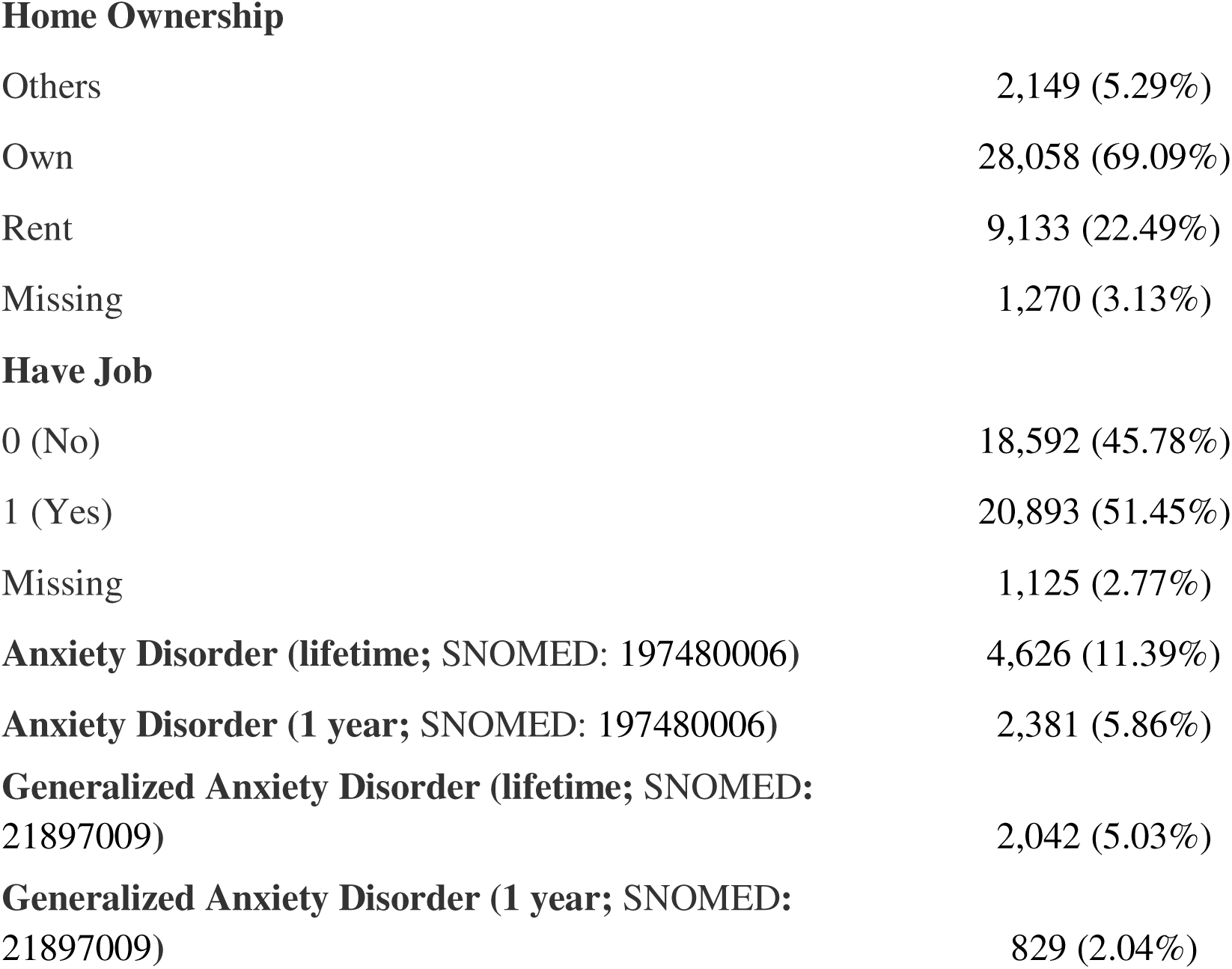
Demographic and Clinical Characteristics of the Study Population. Baseline characteristics of the analytic sample (N=40,610) from the *All of Us* Research Program who completed the July 2020 COPE survey.

| Characteristic | No. (%) of Participants (N = 40,610) |
| --- | --- |
| **Sex** |  |
| Female | 26,151 (64.40%) |
| Male | 13,362 (32.90%) |
| Other | 845 (2.08%) |
| Skipped question | 244 (0.60%) |
| Missing | 8 (0.02%) |
| **Age** |  |
| (18, 44] | 9,207 (22.67%) |
| (44, 64] | 14,146 (34.83%) |
| (64, 90] | 16,987 (41.82%) |
| **Household Income** |  |
| <50k | 10,493 (25.84%) |
| >100k | 14,643 (36.06%) |
| 50k-100k | 11,499 (28.32%) |
| **Education** |  |
| Advanced Degree | 15,141 (37.28%) |
| College Graduate | 12,417 (30.58%) |
| High School or Less | 3,004 (7.40%) |
| Not Answered | 1,042 (2.57%) |
| Partial College | 9,006 (22.18%) |
| **Race** |  |
| Asian | 1,060 (2.61%) |
| Black or African American | 2,132 (5.23%) |
| Others | 3,167 (7.80%) |
| White | 32,980 (81.21%) |
| Missing | 1,280 (3.15%) |
| **Home Ownership** |  |
| Others | 2,149 (5.29%) |
| Own | 28,058 (69.09%) |
| Rent | 9,133 (22.49%) |
| Missing | 1,270 (3.13%) |
| **Have Job** |  |
| 0 (No) | 18,592 (45.78%) |
| 1 (Yes) | 20,893 (51.45%) |
| Missing | 1,125 (2.77%) |
| **Anxiety Disorder (lifetime; SNOMED: 197480006)** | 4,626 (11.39%) |
| **Anxiety Disorder (1 year; SNOMED: 197480006)** | 2,381 (5.86%) |
| **Generalized Anxiety Disorder (lifetime; SNOMED: 21897009)** | 2,042 (5.03%) |
| **Generalized Anxiety Disorder (1 year; SNOMED: 21897009)** | 829 (2.04%) |

### Outcome

Our primary outcome was the sum score of anxiety symptoms measured using the GAD-7 scale (0-21),^13^ during the July 2020 wave of the COPE survey.

### Containment policy exposures

Exposures were seven state-level social distancing policies tracked on a daily basis by the Oxford COVID-19 Government Response Tracker (OxCGRT)^6^: school closings, workplace closings, cancellation of public events, restrictions on gatherings, public transport closures, stay-at-home requirements, and restrictions on internal movement (eTable 1). We also examined the OxCGRT stringency index, a prespecified sum of nine containment indicators. We calculated the average value of each containment policy across each exposure window (see below). Additional exposure measurement details are provided in eMethods.

### Containment policy time windows

The timescale over which containment policies affect anxiety is not known a priori. Exposure windows that are too short may fail to capture effects, while windows that are too long may dilute associations by combining the acute implementation phase with periods of policy adaptation. We therefore evaluated 120 candidate exposure windows defined by varying start dates (March 15 through June 28, 2020, in 7-day increments) and durations (7 to 98 days) to identify the period during which containment policies showed the strongest aggregate associations with July 2020 anxiety symptoms.

To prevent overfitting during window selection, we split the analytic sample into training (n=20,288) and testing (n=20,322) sets using stratified rerandomization. We evaluated 100 candidate partitions stratified by geographic region and GAD-7 severity tertiles, selecting the split with the lowest area under the curve from logistic regression models predicting set membership based on baseline covariates, thereby maximizing covariate balance between sets ^14^ (eFigures 1-5). Regional stratification alone does not guarantee outcome balance because GAD-7 varies within regions; we therefore additionally stratified on GAD-7 tertiles, a coarse discretization of the continuous score, to minimize between-set differences in the outcome distribution. In the training set, we estimated associations using linear regression with cluster-robust standard errors, applied Benjamini-Hochberg false discovery rate correction, and calculated the mean absolute t-statistic across all seven policies to identify the window with the strongest aggregate associations with July 2020 anxiety symptoms (eFigure 6). This procedure identified a 9-week primary exposure window (March 22 through May 23, 2020) where aggregate policy-anxiety associations peaked. We also examined a shorter sensitivity window (April 12 through May 23, 2020; eTable 7). Because the exposure window was selected in the training set based on aggregate association strength, all effect estimates were obtained from the independent testing set to minimize post-selection bias.

### Covariate adjustment

For exposure mixture models, we adjusted for demographic variables (age, sex, self-reported race and ethnicity, birthplace, marital status), socioeconomic indicators (education, employment status, health insurance status), and pre-pandemic anxiety diagnoses derived from EHR data. For the inverse probability weighting model, we incorporated additional pre-pandemic characteristics (e.g., primary language and sexual orientation) and health conditions (e.g., diagnosis of hypertension, obesity, or diabetes in the past year) (see eMethods).

### Statistical analysis

Missing covariate data were addressed using random forest imputation (eMethods). To correct for COPE nonresponse, we fit a propensity score model (logistic regression of survey completion on demographic, socioeconomic, and clinical characteristics) among participants with available EHR data. The inverse of predicted response probabilities served as analytic weights in all subsequent models.

We first examined individual policy effects using linear regression models, adjusting for demographic, socioeconomic, and pre-pandemic anxiety covariates. We then implemented quantile g-computation^10^ to estimate the joint effects of multiple COVID-19 containment policies on the outcome of simultaneously increasing all exposures by one quartile, while deriving empirical weights that reflect each exposure’s proportional contribution to the overall effect. This approach is designed to accommodate correlated exposures, is robust to typical collinearity, and does not require pre-specifying the relative importance of individual policies. We fitted conditional effects models using the average value of each containment policy across the exposure window within the testing set.

All analyses used cluster-robust standard errors at the state level to account for within-state correlation arising from state-level policy exposures and individual-level anxiety outcomes across 50 states. Since mixture weights are data-adaptive parameters,^15^ we determined each policy’s mixture coefficient direction (positive for anxiety-increasing effects, negative for anxiety-decreasing effects) in the training set based on the full seven-policy model. Policies were then partitioned by coefficient direction and re-estimated as separate anxiety-increasing and anxiety-decreasing mixtures in the held-out testing sample to prevent overfitting. As a sensitivity analysis, we applied backward elimination to anxiety-increasing and anxiety-decreasing mixtures separately to identify parsimonious policy subsets (eMethods). We examined whether the effects of containment policies on anxiety varied across age strata, with groups defined based on data availability and prior literature: young adults (18-44 years), middle-aged adults (45-64 years), and older adults (65+ years).

## Results

Participants were predominantly female (64%), with a mean age of 57.80 years (SD=15.76, median=61) (Table 1). Self-reported race/ethnicity distribution included Asian (2.61%), Black (5.23%), White (81.21%), and Other (7.8%) participants. Socioeconomic characteristics included household income above $100,000 for 36%, an advanced degree for 37%, current employment for 52%, and home ownership for 69%. Pre-pandemic anxiety history included 11.39% (n=4626) with a lifetime history of any EHR-documented anxiety disorder (identified via SNOMED code 197480006), with 5.86% diagnosed in the year preceding the pandemic (March 15th, 2019, to March 15th, 2020). For generalized anxiety disorder specifically, 5.03% (n=2,042) of participants had a lifetime GAD diagnosis, and 2.04% (n=829) were diagnosed with GAD in the year preceding the pandemic. Participants reported a mean GAD-7 score of 4.36 (SD=4.77) in July 2020, with 13.3% (n=5,398) reporting moderate-to-severe symptoms (scores 10-21). The distribution of anxiety severity included 61.2% minimal (scores 0-4), 25.5% mild (scores 5-9), 7.9% moderate (scores 10-14), and 5.4% severe (scores 15-21) symptoms (eFigure 2).

### Individual policy associations with anxiety symptoms

We identified a 9-week primary exposure window (March 22 through May 23^rd^, 2020) that showed consistently strong associations across policies and sufficient duration to capture meaningful implementation patterns (eFigures 6-7). Within this window, three *individual* containment policies were associated with lower anxiety symptom scores: cancellation of public events (β = -0.455, 95% CI: -0.813 to -0.096, p = 0.014), restrictions on gathering size (β = - 0.108, 95% CI: -0.183 to -0.033, p=0.006), and stay-at-home requirements (β = -0.634, 95% CI: -1.119 to -0.149, p=0.011). The Oxford Stringency Index^6^, a composite measure of overall policy restrictiveness, also showed a protective association with anxiety symptoms during this window (β = -0.23, 95% CI: -0.44 to -0.03, p = 0.024). No individual policy was independently associated with increased anxiety symptom scores during the primary exposure window. Sensitivity analyses using the full March 15th through June 28th exposure period showed a broadly consistent pattern, though fewer individual policies reached statistical significance (eTable 9).

### Exposure mixture models reveal larger effect sizes than individual policy exposures

We next examined *the joint effects of containment policies* using quantile g-computation, a mixture method that estimates the joint influence of correlated exposures while accounting for their co-occurrence (Figure 1). We partitioned policies by the sign of their mixture coefficient in the joint quantile g-computation model within the training set: policies with positive coefficients formed an “anxiety-increasing” mixture, and policies with negative coefficients formed an “anxiety-decreasing” mixture. In the primary exposure window, the anxiety-increasing mixture comprised school closures, cancellation of public events, workplace closures, and internal travel restrictions; while the anxiety-decreasing mixture included restrictions on gatherings, stay-at-home requirements, and closing public transportation (Table 2).

**Figure 1.**
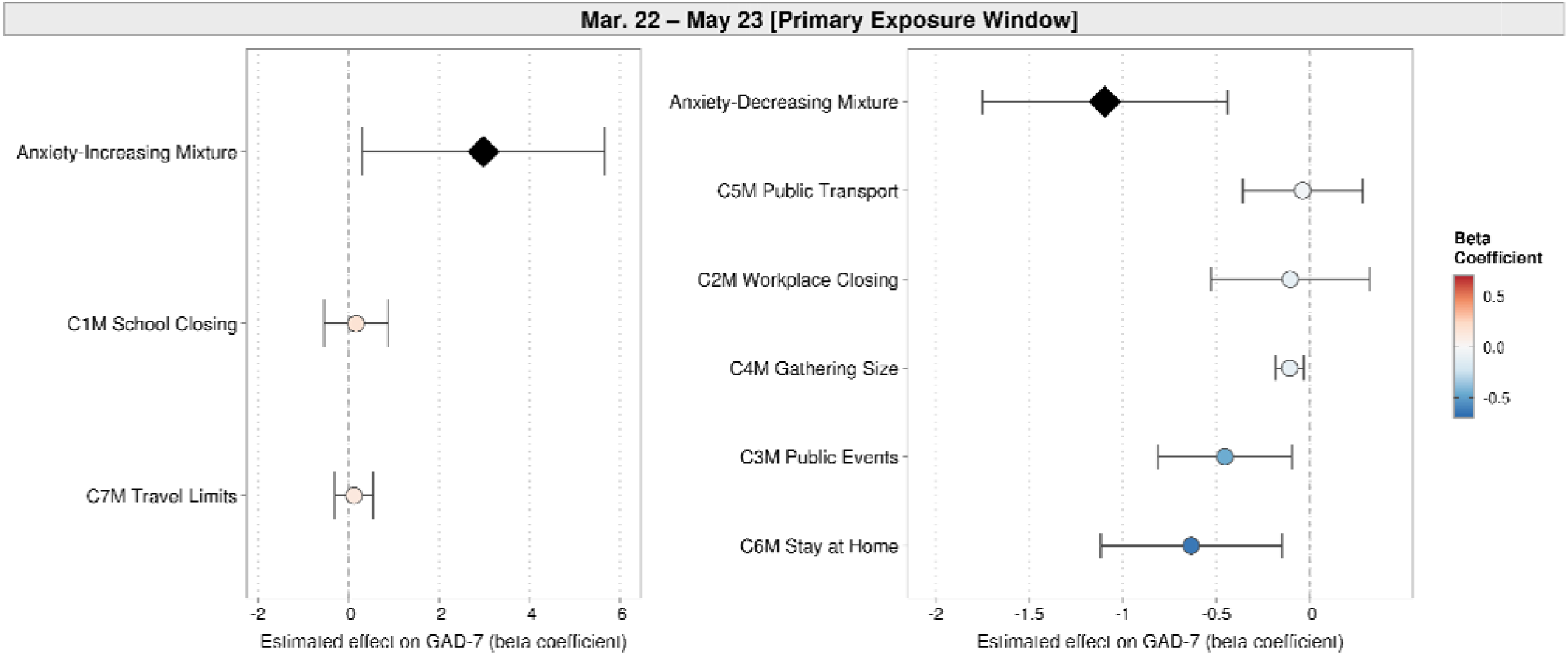
Comparison of single-exposure and mixture effects on anxiety. Forest plots display beta coefficients and 95% confidence intervals comparing individual COVID-19 containment policy effects with combined mixture effects on anxiety symptoms (GAD-7 scale, 0-21) during July 2020. Solid black diamonds indicate statistically significant mixture effects (P ≤ 0.05 after cluster-robust standard error correction), while empty white diamonds indicate non-significant mixture effects. Error bars represent 95% confidence intervals. Covariates included inverse probability treatment weighting of survey response, demographic variables (age, race, marital status), socioeconomic factors (home configuration, education, employment, insurance), and clinical history of pre-pandemic anxiety disorders (any anxiety disorder [SNOMED CT 197480006] and generalized anxiety disorder [SNOMED CT 21897009]) assessed within the past year (March 14, 2019 - March 14, 2020) and lifetime based on electronic health records.

**Table 2.** Individual and mixture policy effects on GAD-7 anxiety symptoms across exposure windows. Associations between seven COVID-19 containment policies and GAD-7 anxiety symptoms during the primary 9-week exposure window (March 22 to May 23, 2020). Three analytical approaches are reported: (1) individual policy associations from separate linear regressions; (2) the Oxford Stringency Index, an equal-weighted composite across all seven policies; and (3) quantile g-computation joint mixture estimates, including the all-policy mixture and the directional partition into anxiety-increasing and anxiety-decreasing mixtures (based on coefficient direction in the training set and re-estimated in the held-out testing set). All estimates were calculated using the testing set (n=20,322). All models were adjusted for demographics, socioeconomic factors, and clinical history of pre-pandemic anxiety disorders, with inverse probability weighting for COPE survey nonresponse. Within each directional mixture, policies are listed in descending order by absolute weight magnitude. Full exposure window (March 15 to June 28, 2020) sensitivity estimates are shown in eTable 9.

|  |  | Exposure time window |  |  |
| --- | --- | --- | --- | --- |
|  |  | Mar 22 - May 23 [Primary Exposure Window] |  |  |
|  |  | Beta | 95% CI | p-value |
| **Individual exposures** |  |  |  |  |
|  | C1M School Closing | 0.168 | (-0.539, 0.875) | 0.635 |
|  | C2M Workplace Closing | -0.105 | (-0.528, 0.319) | 0.622 |
|  | C3M Public Events | -0.455 | (-0.813, -0.096) | **0.014** |
|  | C4M Gathering Size | -0.108 | (-0.183, -0.033) | **0.006** |
|  | C5M Public Transport | -0.039 | (-0.358, 0.281) | 0.809 |
|  | C6M Stay at Home | -0.634 | (-1.119, -0.149) | **0.011** |
|  | C7M Travel Limits | 0.122 | (-0.301, 0.545) | 0.565 |
| **Pre-specified mixture** |  |  |  |  |
|  | Stringency Index (scaled) | -0.234 | (-0.436, -0.032) | **0.024** |
| **Data-adaptive mixtures (weight-partitioned)** |  |  |  |  |
|  | All policies | 1.883 | (-0.509, 4.276) | 0.120 |
|  | Anxiety-increasing mixtures | 2.979 | (0.300, 5.659) | **0.030** |
|  | Selected policies: | C3M, C7M, C2M, C1M |  |  |
|  | Anxiety-decreasing mixtures | -1.096 | (-1.751, -0.440) | **0.002** |
|  | Selected policies: | C6M, C4M, C5M |  |  |

When all seven policies were modeled as a single mixture, we observed no significant associations with anxiety symptoms (β = 1.88, 95% CI: -0.51 to 4.28, p = 0.120), consistent with opposing policy effects offsetting one another in aggregate. However, when policies were partitioned by the direction of their estimated effects (determined in the training set and estimated in the independent testing set), both directional mixtures showed significant associations: the anxiety-increasing mixture was associated with higher anxiety scores (β = 2.98, 95% CI: 0.30 to 5.66, p = 0.030) and the anxiety-decreasing mixture was associated with lower anxiety scores (β = -1.10, 95% CI: -1.75 to -0.44, p = 0.002). These joint mixture effects substantially exceeded the magnitude of any individual policy association, consistent with the accumulation of concurrent policy effects.

### Age stratification reveals opposing policy-anxiety associations between young and older adults

Age-stratified mixtures revealed substantial heterogeneity in the joint effects of containment policies on anxiety symptoms (Figure 2-3, eTable 8). Young adults showed the largest mixture effects of any group during the primary exposure window, with both the anxiety-increasing (β = 8.931, 95% CI: 1.495 to 16.367, p = 0.020) and anxiety-decreasing mixture (β = -2.811, 95% CI: -4.979 to -0.643, p = 0.012) reaching statistical significance. Middle-aged adults (aged 45-64) showed a significant anxiety-increasing mixture effect but no significant anxiety-decreasing mixture effect during the primary exposure window (eTable 8). Older adults showed no significant partitioned effects in the primary exposure window. Further details can be found in Figure 3.

**Figure 2.**
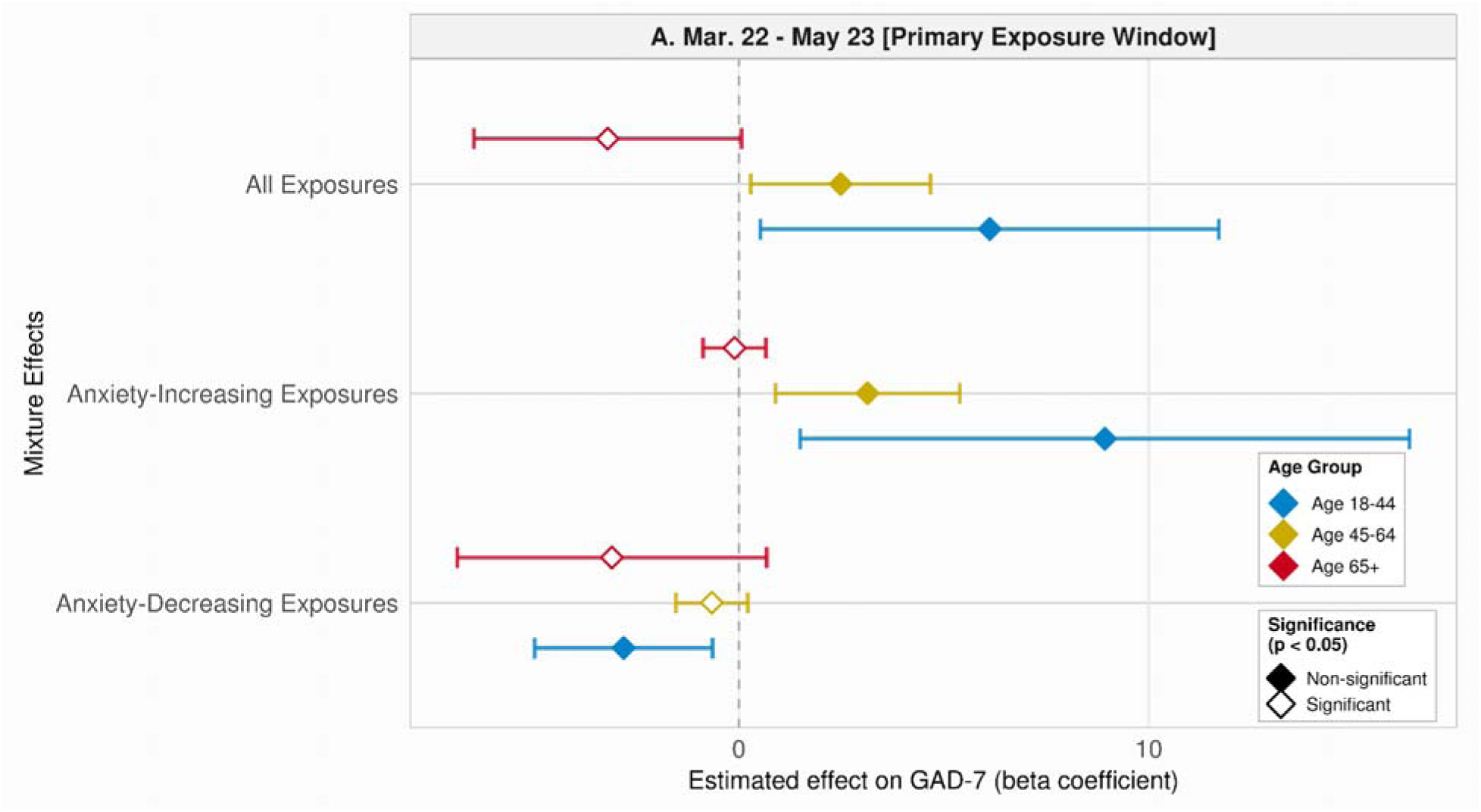
Mixture effects across age strata and exposure windows. Forest plots display beta coefficients and 95% confidence intervals for COVID-19 containment policy mixture effects on anxiety symptoms (GAD-7 scale, 0-21) stratified across age groups within the primary exposure window (March 22-May 23, 2020). Age groups include young adults (18-44 years, n = 9207; blue), middle-aged adults (45-64 years, n = 14,146; yellow), and older adults (65+ years, n = 16,987; red). Filled diamonds indicate statistical significance (P < 0.05) after cluster-robust standard error correction; open diamonds represent non-significant effects. Error bars represent 95% confidence intervals. Covariates and analytic methods are described in Figure 1. Policies were partitioned by their quantile g-computation mixture coefficient directions into anxiety-increasing (positive weights) and anxiety-decreasing (negative weights) groups within the training set, then re-estimated as separate mixture effects in the held-out testing set. Three mixture types were examined, including all policies (joint effect of all seven containment policies), anxiety-increasing policies (policies with positive mixture weights), and anxiety-decreasing policies (policies with negative mixture weights).

**Figure 3.**
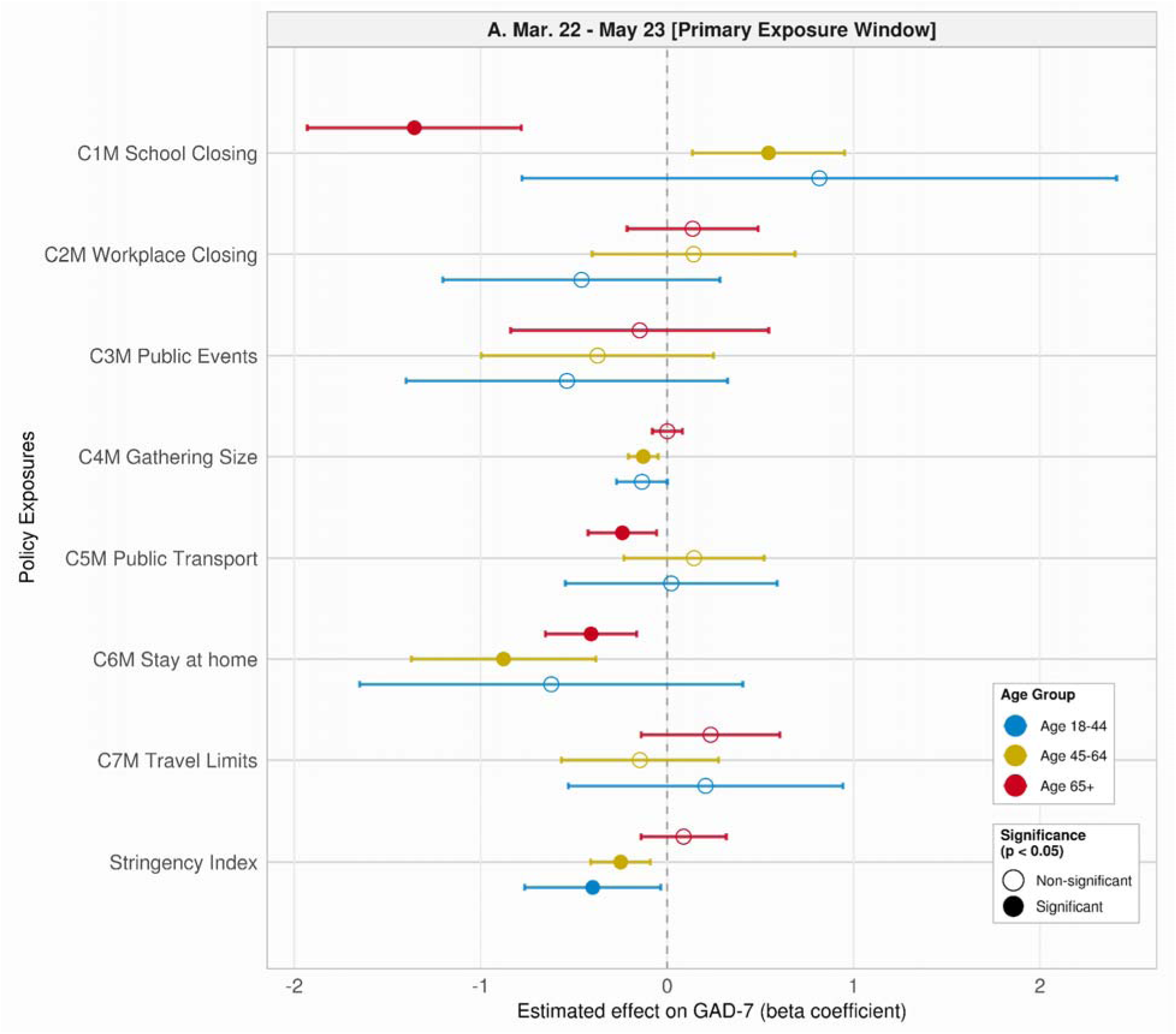
Individual COVID-19 containment policy effects on anxiety symptoms by age strata across exposure windows. Forest plots display beta coefficients and 95% confidence intervals for individual containment policy effects on anxiety symptoms (GAD-7 scale, 0-21) during July 2020, stratified by age group in the primary exposure window (March 22-May 23, 2020). Age groups include young adults (18-44 years, n = 9,207; blue), middle-aged adults (45-64 years, n = 14,146; yellow), and older adults (65+ years, n = 16,987; red). Solid circles indicate statistical significance (P < 0.05) after cluster-robust standard error correction; open circles represent non-significant effects. Error bars represent 95% confidence intervals. The vertical dashed line represents null effect (β = 0). Each policy was estimated via separate linear regression with covariate adjustment as described in Figure 1.

## Discussion

During the COVID-19 pandemic, governments implemented containment policies to reduce viral transmission, yet these measures carried potential costs for population mental health. Knowing which policy combinations increased or decreased anxiety is essential for weighing transmission control against mental health costs in future pandemics. Prior work has largely relied on unweighted aggregate indices, such as the OxCGRT Stringency Index, which assigns equal weight to each policy and cannot distinguish the mental health contribution of any single policy. Studies that instead examined individual policies in isolation have shown inconsistent findings, likely because policies were implemented simultaneously and their effects depend on which other measures were in place concurrently. Neither approach captures how different combinations of containment policies jointly affect mental health. We addressed this gap using quantile g-computation, an exposure mixture method that empirically weights distinct policy contributions within a joint model, to estimate how specific policy combinations influenced anxiety symptoms in a large, nationwide cohort from the *All of Us* Research Program. As such, ours is the first study to evaluate the impact of specific combinations of containment policies.

When all seven containment policies were modeled jointly during the 9-week primary exposure window (March 22 through May 23, 2020), the estimated joint effect on GAD-7 anxiety scores was a non-significant increase of approximately 2 points on a 21-point scale. This magnitude may have population-level relevance if confirmed in larger samples. Large longitudinal studies using the OxCGRT Stringency Index have linked higher stringency to increased anxiety and depressive symptoms across European cohorts^16–17^, although a dose-response meta-analysis found these associations attenuated after adjustment for cumulative COVID-19 cases.^18^

At the individual-policy level, our results showed that three of the seven policies were significantly protective during the primary window (cancellation of public events, restrictions on gathering size, and stay-at-home requirements), while the remaining four showed no significant individual association. Our findings diverged from several prior studies. Stay-at-home requirements were associated with increased anxiety in a Canadian sample,^19^ and a meta-regression of 60 studies identified public transport closures as the only policy consistently associated with anxiety.^20^ Mendez-Lopez and colleagues,^9^ analyzing European older adults, observed that stay-at-home orders, gathering restrictions, and public transport closures were each associated with worsening mental health, whereas we found gathering restrictions and stay-at-home requirements protective both individually and within the anxiety-decreasing mixture described below. Other work has reported protective effects consistent with our findings, including improved mental well-being associated with workplace closures among non-key workers and middle-aged adults.^21^ Several differences in study design likely account for these divergences, including outcome measurement, sample composition, and exposure timing. Mendez-Lopez and colleagues,^9^ for example, relied on retrospective self-report of perceived worsening of anxiety, which captures a different construct than current symptom severity assessed cross-sectionally using the GAD-7^13^, and their older-adult-only sample differs from our analytic sample, which spans the full adult age range. These design differences offer one interpretation; the observed heterogeneity is also consistent with policies whose joint effects counteract one another, a possibility that neither single-policy estimates nor the all-policy aggregate mixture can fully address.

When the same seven policies were partitioned by the sign of their mixture coefficient in the training set and re-estimated in the testing set, both directional mixtures showed significant associations with magnitudes substantially exceeding any individual policy. The anxiety-increasing combination (school closures, cancellations of public events, workplace closures, and internal travel restrictions) was associated with a 3-point higher GAD-7 score (β = 2.98, 95% CI: 0.30 to 5.66); the anxiety-decreasing mixture (gathering restrictions, public transport closures, and stay-at-home requirements) was associated with a 1-point lower score (β = -1.10, 95% CI: - 1.75 to -0.44). The 4-point minimally clinically important difference (MCID) for the GAD-7^22^ is defined for within-individual change; a population-level mean shift of 3 points is smaller than the MCID threshold but would, in a sample where roughly a quarter reported mild subthreshold symptoms (GAD-7 scores 5-9), meaningfully increase the proportion in the moderate-to-severe range (GAD-7 scores 10-21).

Supplementary analyses extending the exposure window through June 28, 2020, show attenuation of both directional associations over time: the anxiety-increasing and anxiety-decreasing mixtures showed smaller coefficient magnitudes in the longer window, and the anxiety-decreasing mixture was no longer significant. Prior studies align with the same time-dependent decay in effects: longitudinal studies show that anxiety symptoms peaked at the start of the pandemic^22^ and declined as the first lockdown progressed and as restrictions continued, reaching their lowest prevalence by early July 2020.^24^ Similarly, a recent target trial emulation in nationally representative U.S. data found that lifting full lockdown temporarily reduced psychological distress, with effects becoming negligible within two months.^25^ One interpretation is that the early window captured an acute phase in which restrictions affected a population still adapting to a novel virus threat, with habituation to sustained restrictions and behavioral adjustment dampening policy-anxiety associations as the exposure window lengthened. An alternative, non-exclusive explanation is that time-varying outbreak severity, including case counts and policy lifting itself, confounded exposure and outcome trajectories over a longer time horizon.

Young adults (age 18-44) showed the largest anxiety-increasing mixture association of any age group, with a GAD-7 increase of β = 8.93 points (95% CI: 1.50 to 16.37). While the point estimate exceeded the 4-point clinically meaningful threshold,^18^ the confidence interval spanned 71% of the 21-point scale, indicating substantial imprecision. Replication in larger samples is needed before drawing clinical conclusions from this subgroup estimate. Prior evidence has shown that pandemic restrictions disproportionately affected the mental health of young adults relative to older adults.^26–28^ Plausible contributors to elevated young-adult vulnerability include greater reliance on workplace environments for social connection,^29^ higher likelihood of experiencing employment disruption from workplace closures,^30^ and potential caregiving responsibilities that increase susceptibility to the mental health impacts of containment measures.^31^ School closures showed associations in opposite directions across age groups: the policy was associated with increased anxiety in young and middle-aged adults but decreased anxiety in older adults (eTable 8), consistent with educational disruptions or increased childcare burden in younger and middle-aged adults^26^ and reduced perceived exposure risk among older adults at higher COVID-19 mortality risk.^27^

Our analytical framework incorporated several design features to mitigate common methodological limitations. We used stratified rerandomization to create balanced training and testing sets, estimated exposure windows empirically to avoid arbitrary time-period selection, applied inverse probability weighting to address COPE survey nonresponse bias, and used cluster-robust standard errors at the state level to account for within-state correlation in policy exposures and anxiety outcomes.

## Limitations

This study has several limitations. First, our analysis relied on participants enrolled in the *All of Us* Research Program who completed the COPE survey and was restricted to the first months of the pandemic. While inverse probability weighting corrected for COPE survey nonresponse within the *All of Us* source population, results may not be generalizable to other populations or exposure periods. Second, policy stringency is endogenous to outbreak severity: states with higher infection and mortality rates tended to adopt stricter containment measures. Associations between high-stringency policies and anxiety symptoms therefore likely reflect both the containment response and the perceived threat of the virus, rather than the policy effect in isolation. Because the relevant confounding is time-varying (the within-state outbreak trajectory unfolded over the exposure window), our cross-sectional design cannot separate these two sources. The state-level fixed effects we adjusted for capture only time-invariant characteristics, and a single cross-sectional case-count adjustment cannot capture exposure-window dynamics. Future studies could separate these contributions using repeated-measures data (daily state-level case counts and deaths) analyzed with longitudinal methods such as marginal structural models exploiting policy-timing variation across states. Third, our exposure measures captured the enactment of policies, not public adherence or local enforcement, which could mediate or confound the observed associations. Averaging policy levels across each exposure window also captures overall intensity but masks within-window variation in the timing of implementation and rate of policy change; restricting the primary analysis to a narrow 9-week window and conducting a shorter sensitivity-window analysis (eTable 7) partly mitigates though does not eliminate this concern. Fourth, because participants did not complete the GAD-7 before the pandemic, our outcome reflects the cross-sectional level of anxiety symptoms in July 2020 rather than within-person change. Adjustment for EHR-documented pre-pandemic anxiety diagnoses partially accounts for pre-existing anxiety burden, but does not capture undiagnosed cases, subclinical pre-pandemic symptoms, or pre-pandemic symptom severity. Fifth, while we examined age-related moderation, other potential moderators, such as employment status, caregiving responsibilities, or pre-existing mental health conditions were not examined and warrant future investigation. Sixth, our analysis assumed linearity and additivity of exposure effects, which may not capture more complex patterns such as exposure interactions, and future work using alternative mixture methods that relax these assumptions^32^ may provide useful comparisons.

## Conclusion

The impact of pandemic-related containment policies on mental health outcomes has important implications for future pandemic preparedness. We found that specific sets of COVID-19 containment policies produced substantially larger joint associations with anxiety than analyses of individual policies would suggest. School closures, cancellation of public events, workplace closures, and travel limits jointly showed anxiety-increasing associations, while public transport closures, gathering restrictions, and stay-at-home requirements jointly showed anxiety-decreasing associations. Younger adults experienced substantially higher anxiety compared to older age groups, suggesting that future pandemic preparedness should consider age-specific vulnerability patterns when designing containment strategies. These results suggest that containment policies should be evaluated jointly rather than in isolation, with attention to their combined effects that may be anxiety-increasing or anxiety-decreasing depending on the specific policy combination.

## Data Availability

This study used data from the All of Us Research Program's Controlled Tier Dataset (version v7), available to authorized researchers on the Researcher Workbench (https://www.researchallofus.org/). Individual-level All of Us data are not publicly available; access requires an institutional Data Use and Registration Agreement and completion of the program's registration and training requirements. State-level COVID-19 containment policy data are publicly available from the Oxford COVID-19 Government Response Tracker (OxCGRT), as cited in the manuscript.

## Acknowledgments

This work was supported in part by NIMH RF1MH134638 (to JWS). Dr. Brunoni is the recipient of the Sherry Gold Knopf Crasilneck Distinguished Chair in Psychiatry, in Honor of Mollie and Murray Gold, and of the Dear Scholar Award of the University of Texas Southwestern Medical Center.

## References

1. COVID-19 Mental Disorders Collaborators. Global prevalence and burden of depressive and anxiety disorders in 204 countries and territories in 2020 due to the COVID-19 pandemic. Lancet. 2021;398(10312):1700-1712.

2. Delpino FM, da Silva CN, Jerônimo JS, et al. Prevalence of anxiety during the COVID-19 pandemic: A systematic review and meta-analysis of over 2 million people. J Affect Disord. 2022;318:272-282.

3. Great Barrington Declaration. Great Barrington Declaration. April 13, 2020. Accessed May 11, 2026. https://web.archive.org/web/20260509153955/https://gbdeclaration.org/

4. Alwan NA, Burgess RA, Ashworth S, et al. Scientific consensus on the COVID-19 pandemic: we need to act now. Lancet. 2020;396(10260):e71–e72.

5. Penninx BWJH, Benros ME, Klein RS, Vinkers CH. How COVID-19 shaped mental health: from infection to pandemic effects. Nat Med. 2022;28(10):2027–2037.

6. Hale T, Angrist N, Goldszmidt R, et al. A global panel database of pandemic policies (Oxford COVID-19 Government Response Tracker). Nat Hum Behav. 2021;5(4):529–538.

7. Aknin LB, Andretti B, Goldszmidt R, et al. Policy stringency and mental health during the COVID-19 pandemic: a longitudinal analysis of data from 15 countries. Lancet Public Health. 2022;7(5):e417–e426.

8. Varga TV, Bu F, Dissing AS, et al. Loneliness, worries, anxiety, and precautionary behaviours in response to the COVID-19 pandemic: A longitudinal analysis of 200,000 Western and Northern Europeans. Lancet Reg Health Eur. 2021;2:100020.

9. Mendez-Lopez A, Stuckler D, McKee M, Semenza JC, Lazarus JV. The mental health crisis during the COVID-19 pandemic in older adults and the role of physical distancing interventions and social protection measures in 26 European countries. SSM Popul Health. 2022;17:101017.

10. Keil AP, Buckley JP, O’Brien KM, Ferguson KK, Zhao S, White AJ. A Quantile-Based g-Computation Approach to Addressing the Effects of Exposure Mixtures. Environ Health Perspect. 2020;128(4):47004.

11. 11. All of Us Research Program Investigators, Denny JC, Rutter JL, et al. The “All of Us” Research Program. N Engl J Med. 2019;381(7):668-676.

12. Schulkey CE, Litwin TR, Ellsworth G, et al. Design and Implementation of the All of Us Research Program COVID-19 Participant Experience (COPE) Survey. Am J Epidemiol. 2023;192(6):972–986.

13. Spitzer RL, Kroenke K, Williams JBW, Löwe B. A brief measure for assessing generalized anxiety disorder: the GAD-7. Arch Intern Med. 2006;166(10):1092–1097.

14. Morgan KL, Rubin DB. Rerandomization to Balance Tiers of Covariates. J Am Stat Assoc. 2015;110(512):1412–1421.

15. Hubbard AE, Kherad-Pajouh S, van der Laan MJ. Statistical Inference for Data Adaptive Target Parameters. Int J Biostat. 2016;12(1):3–19.

16. Bu F, Steptoe A, Fancourt D. Depressive and anxiety symptoms in adults during the COVID-19 pandemic in England: A panel data analysis over 2 years. PLOS Medicine. 2023;20(4):e1004144.

17. Hajek A, Neumann-Böhme S, Sabat I, et al. Depression and anxiety in later COVID-19 waves across Europe: New evidence from the European COvid Survey (ECOS). Psychiatry Res. 2022;317:114902.

18. Salanti G, Peter N, Tonia T, et al. The Impact of the COVID-19 Pandemic and Associated Control Measures on the Mental Health of the General Population : A Systematic Review and Dose-Response Meta-analysis. Ann Intern Med. 2022;175(11):1560–1571.

19. Plett D, Pechlivanoglou P, Coyte PC. The impact of provincial lockdown policies and COVID-19 case and mortality rates on anxiety in Canada. Psychiatry Clin Neurosci. 2022;76(9):468–474.

20. Castaldelli-Maia JM, Marziali ME, Lu Z, Martins SS. Investigating the effect of national government physical distancing measures on depression and anxiety during the COVID-19 pandemic through meta-analysis and meta-regression. Psychol Med. 2021;51(6):881–893.

21. Toffolutti V, Plach S, Maksimovic T, et al. The association between COVID-19 policy responses and mental well-being: Evidence from 28 European countries. Soc Sci Med. 2022;301:114906.

22. Anne Toussaint, Paul Hüsing, Antje Gumz, Katja Wingenfeld, Martin Härter, Elisabeth Schramm, Bernd Löwe. Sensitivity to change and minimal clinically important difference of the 7-item Generalized Anxiety Disorder Questionnaire (GAD-7). Journal of Affective Disorders. 2020;265:395–401.

23. Salanti G, Peter NL, Tonia T, et al. Changes in the prevalence of mental health problems during the first year of the pandemic: a systematic review and dose-response meta-analysis. BMJ Ment Health. 2024;27(1). doi:10.1136/bmjment-2024-301018

24. Ori APS, Wieling M, Lifelines Corona Research Initiative, van Loo HM. Longitudinal analyses of depression, anxiety, and suicidal ideation highlight greater prevalence in the northern Dutch population during the COVID-19 lockdowns. J Affect Disord. 2023;323:62-70.

25. Cudic M, de la Hoz JF, Dall’Aglio L, et al. Would Lifting Versus Maintaining COVID-19 Containment Policies Have Reduced Psychological Distress in the US? medRxiv. Published online March 9, 2026:2026.03.06.26347802. doi:10.64898/2026.03.06.26347802

26. Sojli E, Tham WW, Bryant R, McAleer M. COVID-19 restrictions and age-specific mental health-U.S. probability-based panel evidence. Transl Psychiatry. 2021;11(1):418.

27. Nichols E, Petrosyan S, Khobragade P, et al. Trajectories and correlates of poor mental health in India over the course of the COVID-19 pandemic: a nationwide survey. BMJ Glob Health. 2024;9(1). doi:10.1136/bmjgh-2023-013365

28. Koch M, Park S. Do government responses impact the relationship between age, gender and psychological distress during the COVID-19 pandemic? A comparison across 27 European countries. Soc Sci Med. 2022;292:114583.

29. Breetzke J, Wild EM. Social connections at work and mental health during the first wave of the COVID-19 pandemic: Evidence from employees in Germany. PLoS One. 2022;17(6):e0264602.

30. Li L, Serido J, Vosylis R, et al. Employment Disruption and Wellbeing Among Young Adults: A Cross-National Study of Perceived Impact of the COVID-19 Lockdown. J Happiness Stud. 2023;24(3):991–1012.

31. Wiedemann A, Stochl J, Neufeld SAS, et al. The impact of the initial COVID-19 outbreak on young adults’ mental health: a longitudinal study of risk and resilience factors. Sci Rep. 2022;12(1):16659.

32. McCoy D, Hubbard A, Van der Laan M. CVtreeMLE: Efficient Estimation of Mixed Exposures using Data Adaptive Decision Trees and Cross-Validated Targeted Maximum Likelihood Estimation in R. J Open Source Softw. 2023;8(82). doi:10.21105/joss.04181

