## Supplementary Material for "Joint Effects of Early Pandemic Containment Policies on Anxiety in the United States"

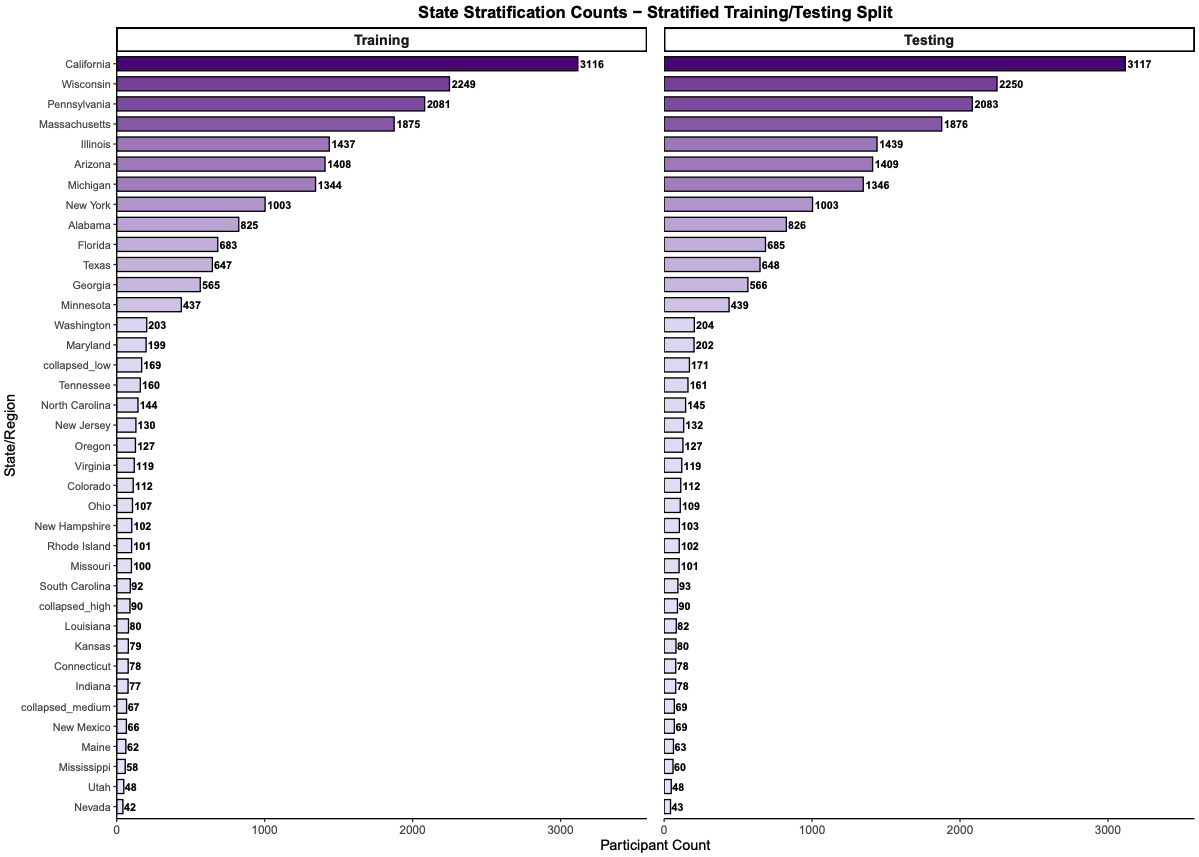

**eFigure 1. Stratification of geographic region between training and testing samples.** Distribution of participants across geographic regions between training (n=20,288) and testing (20,322) sets following stratified sampling of geographic regions and GAD-7 quantile bins, and rerandomization to further minimize imbalances in patient characteristics. To ensure adequate sample sizes for stratification, 15 states with small strata were collapsed into three groups based on state-level COVID-19 policy stringency index tertiles: collapsed low (stringency < 57.59; n=340), collapsed medium (stringency 57.59-64.81; n = 136), and collapsed high (stringency > 64.81; n=180). Collapsed states included Alaska, Arkansas, Delaware, Hawaii, Idaho, Iowa, Kentucky, Montana, Nebraska, North Dakota, Oklahoma, South Dakota, Vermont, West Virginia, and Wyoming. The remaining 35 states maintained individual representation due to sufficient sample sizes across all strata.

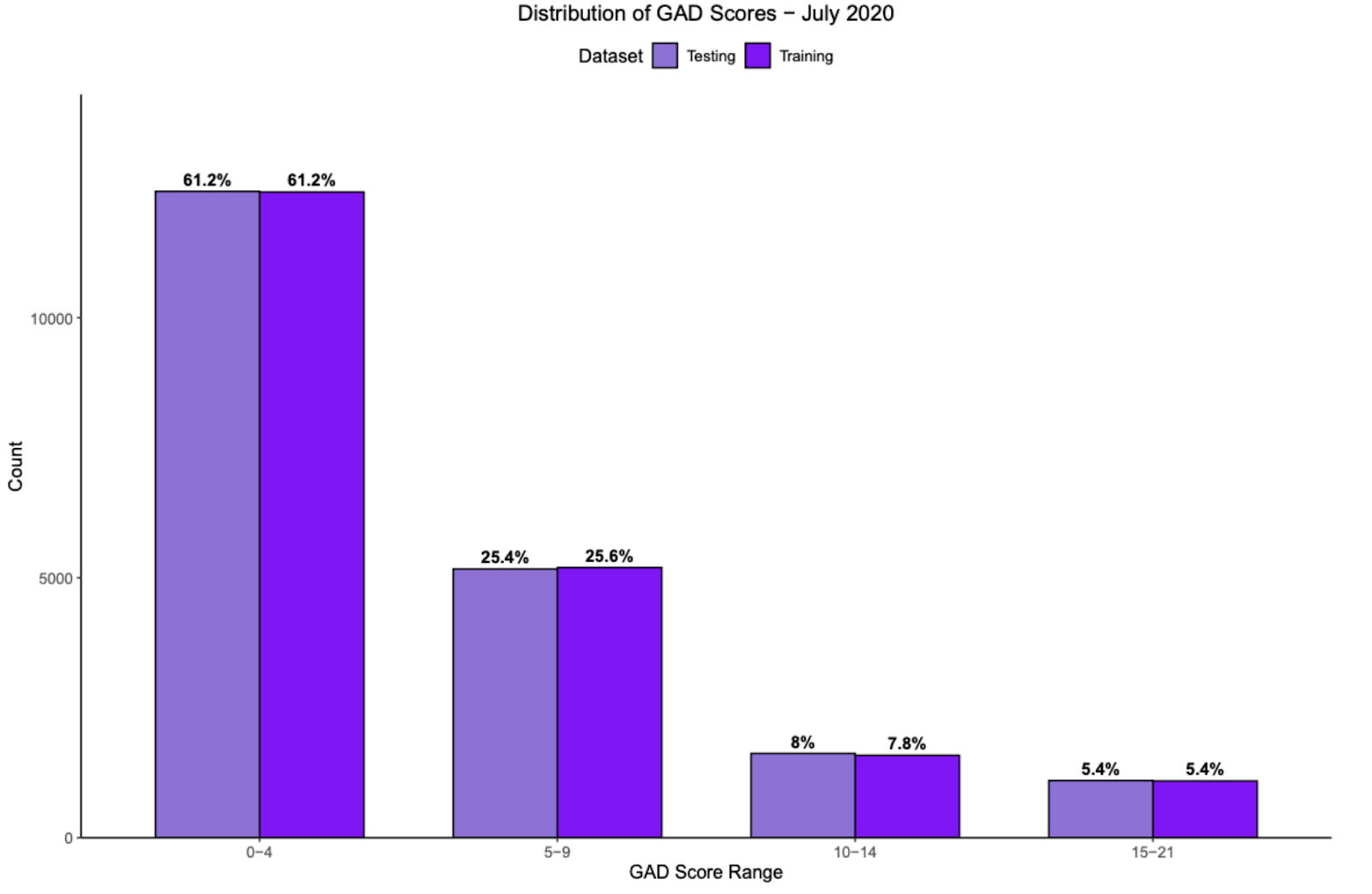

**eFigure 2. Distribution of GAD-7 anxiety scores during the July 2020 outcome window.** Distribution of GAD-7 anxiety scores in July 2020 across training (dark purple) and testing (light purple) sets following stratified sampling and rerandomization. For stratified sampling, GAD-7 scores were divided into tertiles: None (0-1; n=10,41), Minimal (1-5; n=14,006), and Mild/Severe (5-21; n=15,763), which were used in conjunction with geographic state groupings to ensure balanced randomization across training (n=20,288) and testing (20,322) sets. The distributions show balanced partitions across all clinical severity ranges, with differences between training and testing sets of less than 0.3 percentage points (0-4: 61.2% vs 61.2%; 5-9: 25.4% vs 25.6%; 10-14: 8% vs 7.8%; 15-21: 5.4% vs 5.4%). Within quantile g-computation exposure mixture models, GAD-7 scores were analyzed on their original continuous scale (0-21) to preserve full variability in anxiety symptom severity. The tertile stratification was used exclusively for balanced randomization during data partitioning, not for outcome analysis. This stratification approach, combined with geographic state groupings, ensured comparable distributions of both exposure variables (state-level containment policies) and outcome severity between training and testing sets.

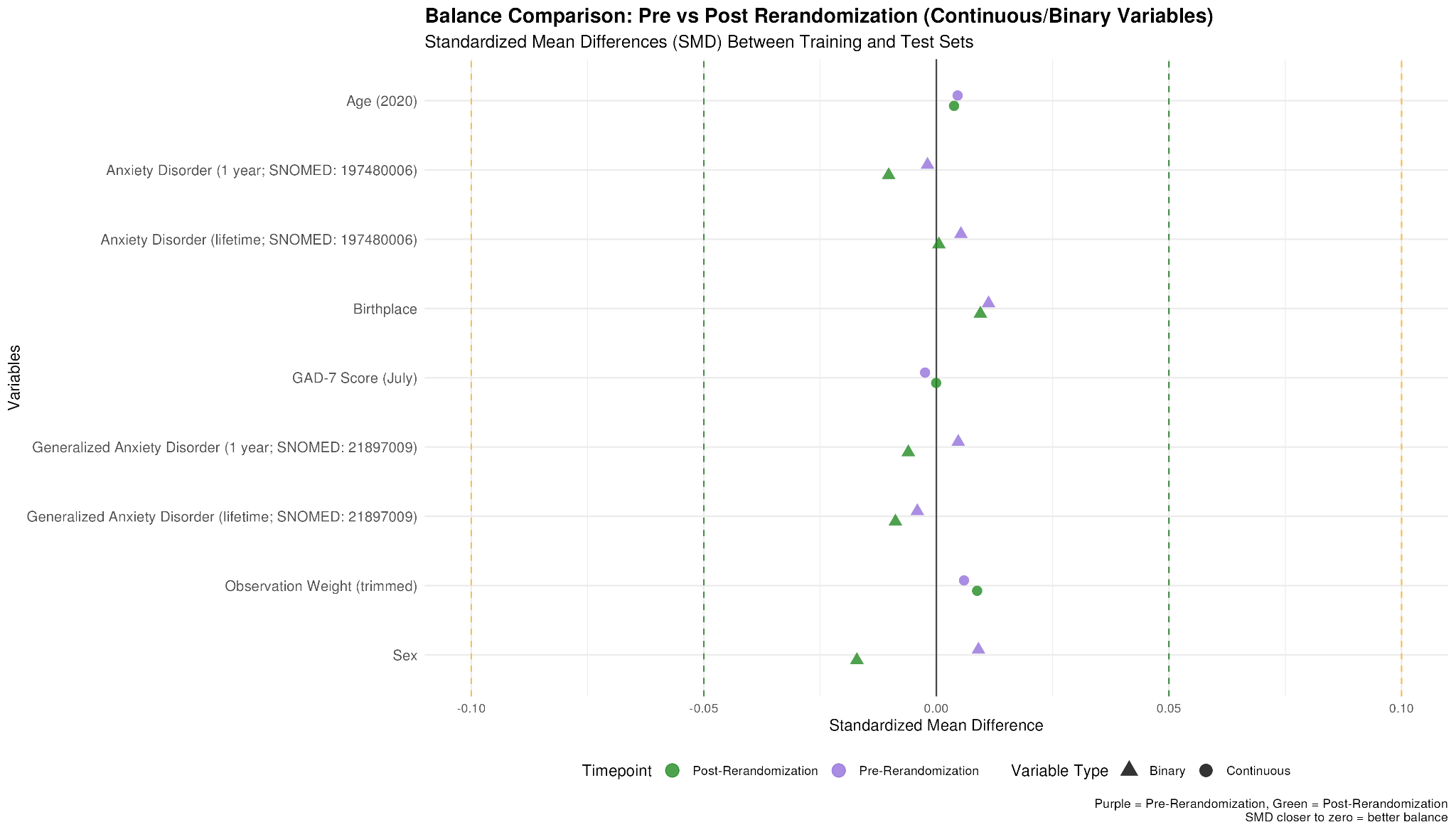

**eFigure 3. Standardized mean differences for adjustment variables between training and testing sets before and after rerandomization.** Standardized mean differences (SMD) for adjustment variables used in quantile g-computation exposure mixture models, comparing training and testing sets before (purple) and after (green) rerandomization. The SMD quantifies the difference in means between groups scaled by the pooled standard deviation, providing a standardized measure of covariate balance independent of variable units or scales. Values below 0.05 indicate excellent balance, while values above 0.1 suggest imbalance that could bias model performance comparisons. All observed SMD values were well below the excellent balance threshold (range: 0.002-0.011), indicating successful balance of model adjustment variables between training and testing partitions. Post-rerandomization SMD values showed marginal improvements for most adjustment variables, providing justification for the iterative rerandomization procedure for optimizing training-testing partition balance.

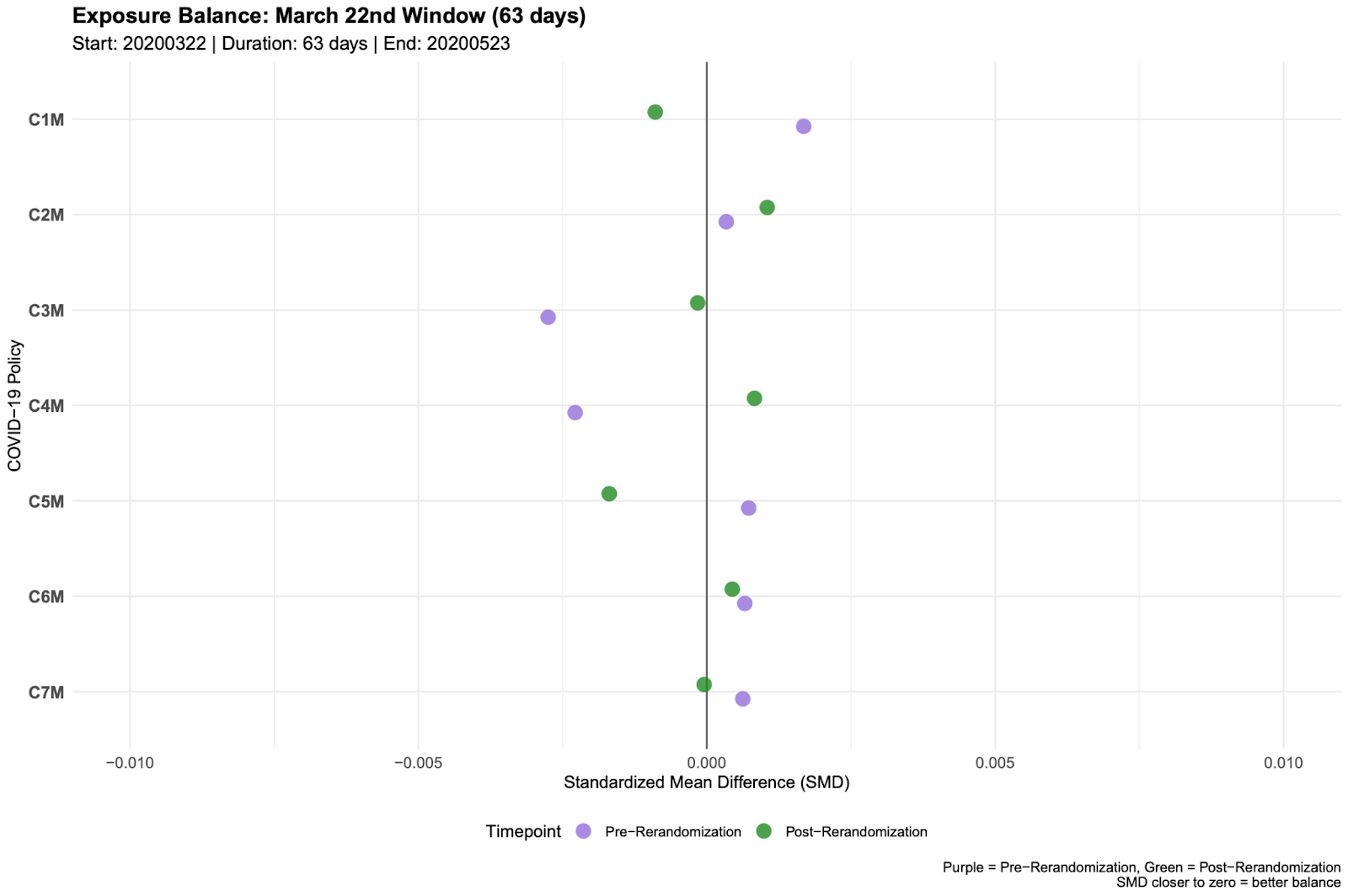

**eFigure 4. Exposure balance assessment for COVID-19 containment policies between training and testing sets across selected exposure windows.** Standardized mean differences (SMD) for COVID-19 containment policy exposures between training and testing sets before (purple) and after (green) rerandomization across the primary exposure window from March 22nd-May 23rd (63 days). Policy exposures include school closures (C1M), workplace closures (C2M), cancellation of public events (C3M), restrictions on gatherings (C4M), public transportation closures (C5M), stay-at-home requirements (C6M), and internal travel limits (C7M). The SMD quantifies the difference in mean exposure levels between training and testing sets scaled by the pooled standard deviation, providing a standardized measure of exposure balance independent of policy-specific scales. Values closer to zero indicate better balance between groups, with SMD values below 0.05 generally considered excellent balance. All observed SMD values were well below 0.005 across all policies and exposure windows, both before and after rerandomization, indicating excellent balance in COVID-19 containment policy exposures between training and testing sets. Kolmogorov-Smirnov tests comparing the full distributional shapes of policy exposures between training and testing sets (eTable 3) further confirmed the absence of meaningful distributional differences across all exposure windows.

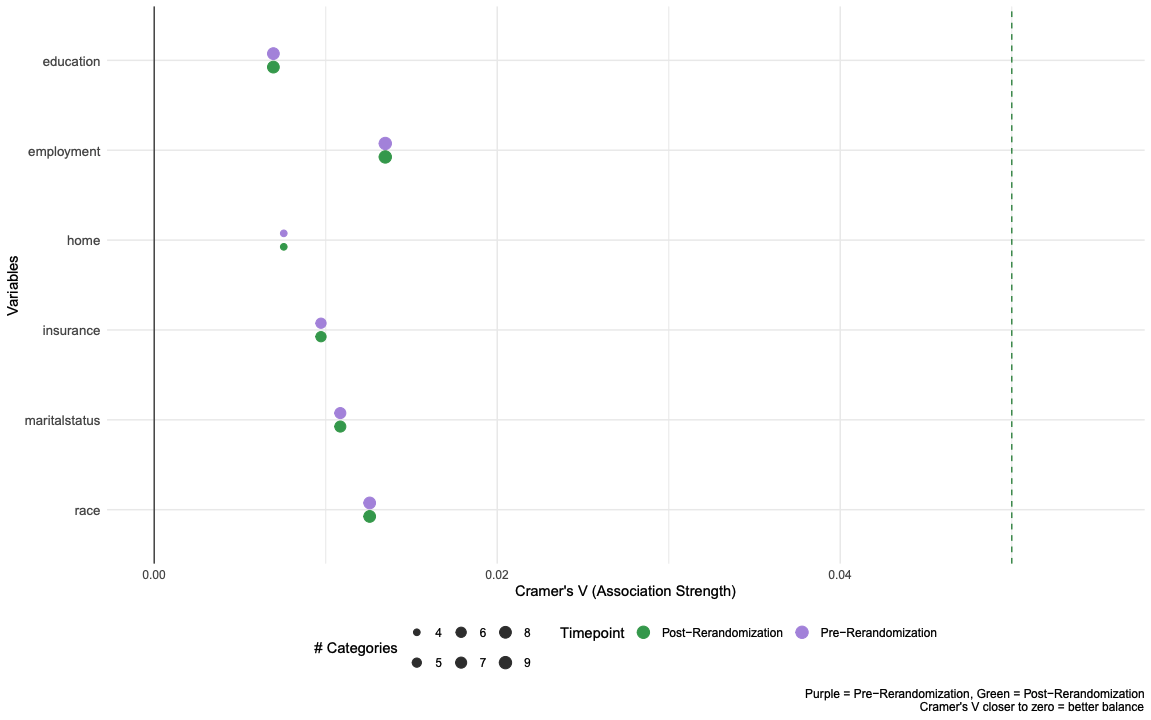

**eFigure 5. Categorical variable balance assessment using cramer’s v between training and testing sets before and after rerandomization.** Cramer’s V statistics for categorical adjustment variables used in quantile g-computation exposure mixture models, comparing training and testing sets before (purple) and after (green) rerandomization. Cramer’s V measures the strength of association between two categorical variables, derived from chi-square statistics and normalized for sample size and degrees of freedom to range from 0 (no association) to 1 (perfect association). Values approaching zero indicate excellent balance between groups, while values above 0.1 suggest small effects that may indicate meaningful imbalance. All observed Cramer’s V values were well below 0.02 (range: 0.007-0.014), indicating excellent balance across all categorical adjustment variables between training and testing partitions. The minimal differences between pre- and post- rerandomization values demonstrate that both approaches achieved adequate categorical balance.

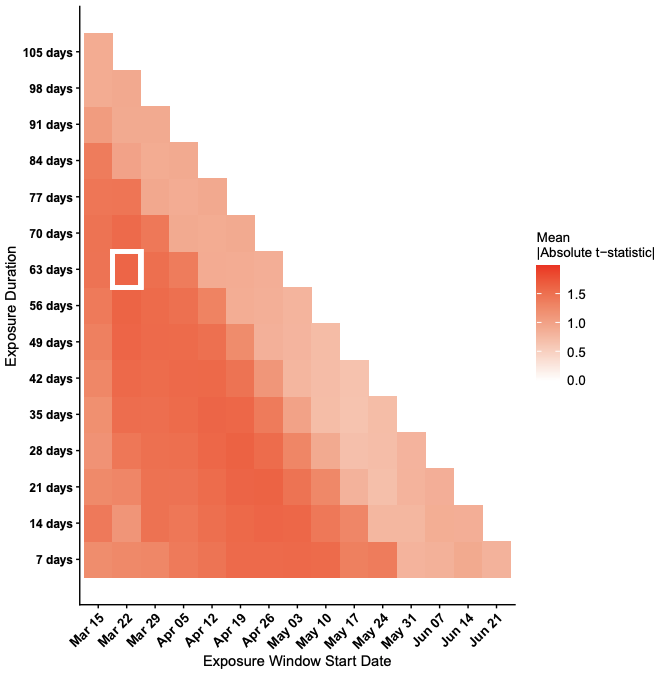

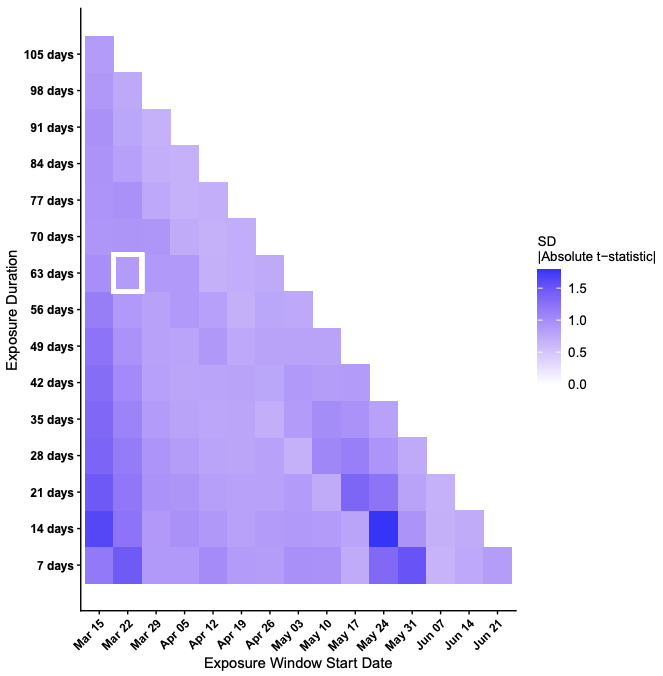

**eFigure 6. Identification of exposure windows with strongest containment policy-anxiety association.** Using the training set (n=20,288), this figure displays results from evaluating 840 policy-window combinations (7 policies x 120 windows) to identify periods where COVID-19 containment policies showed the strongest associations with GAD-7 anxiety symptoms measured in July 2020. Linear regression models were performed for each combination with cluster-robust standard error correction at the state level to account for within-state correlation in outcomes arising from shared state-level policy exposures, where exposure windows varied in 7-day increments from March 15th through June 21st, 2020, and the final window ended June 28th, 2020. Panel A displays the mean absolute t-statistics averaged across all seven policies for each exposure window, with darker red indicating periods where policies collectively demonstrated stronger associations with anxiety outcomes. Panel B displays the standard deviation of absolute t-statistics across policies within each window, with darker blue indicating greater between-policy variability in association strengths. The white box outlines the selected data-adaptive exposure window (March 22nd-May 23rd, 2020) used in subsequent mixture analyses, chosen based on consistently strong associations across policies and sufficient duration to capture meaningful policy implementation patterns.

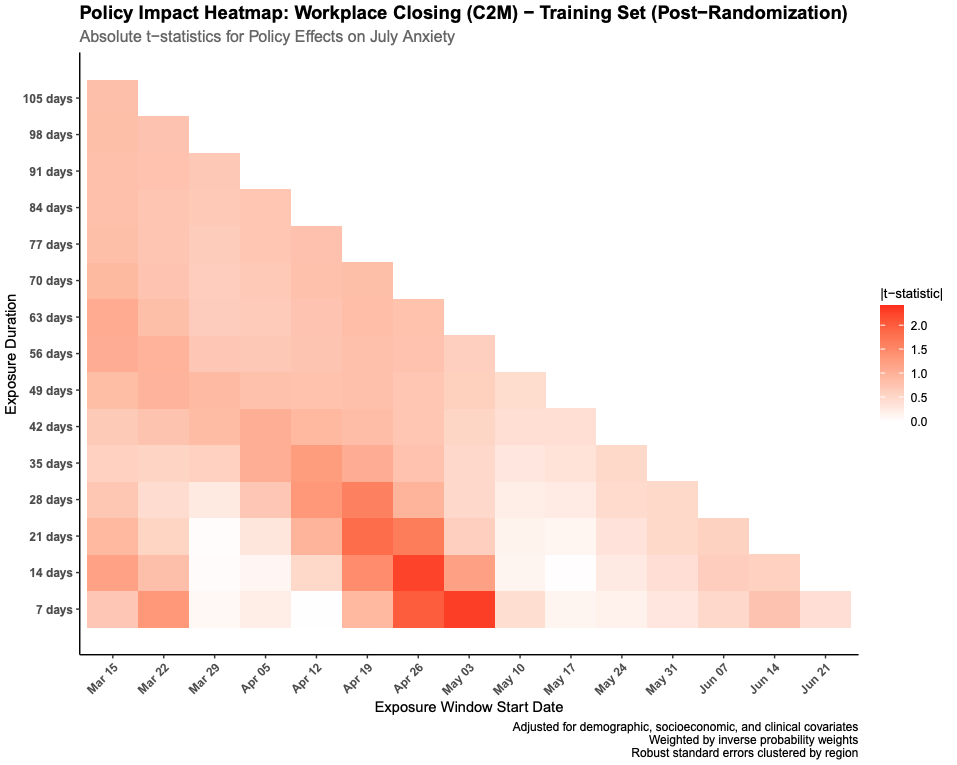

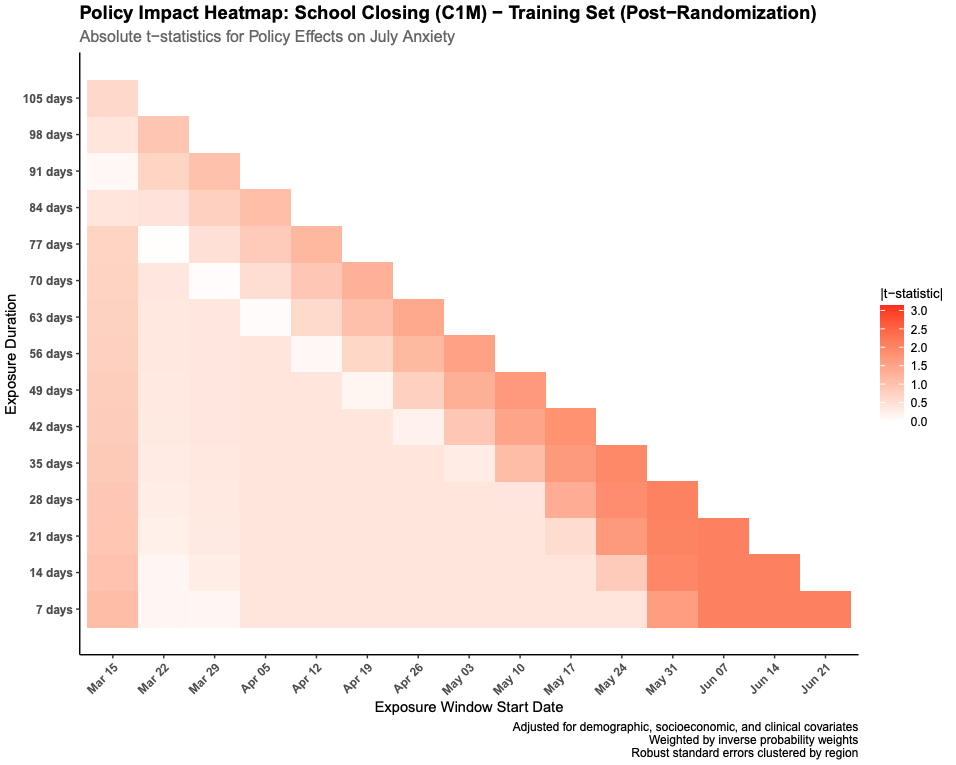

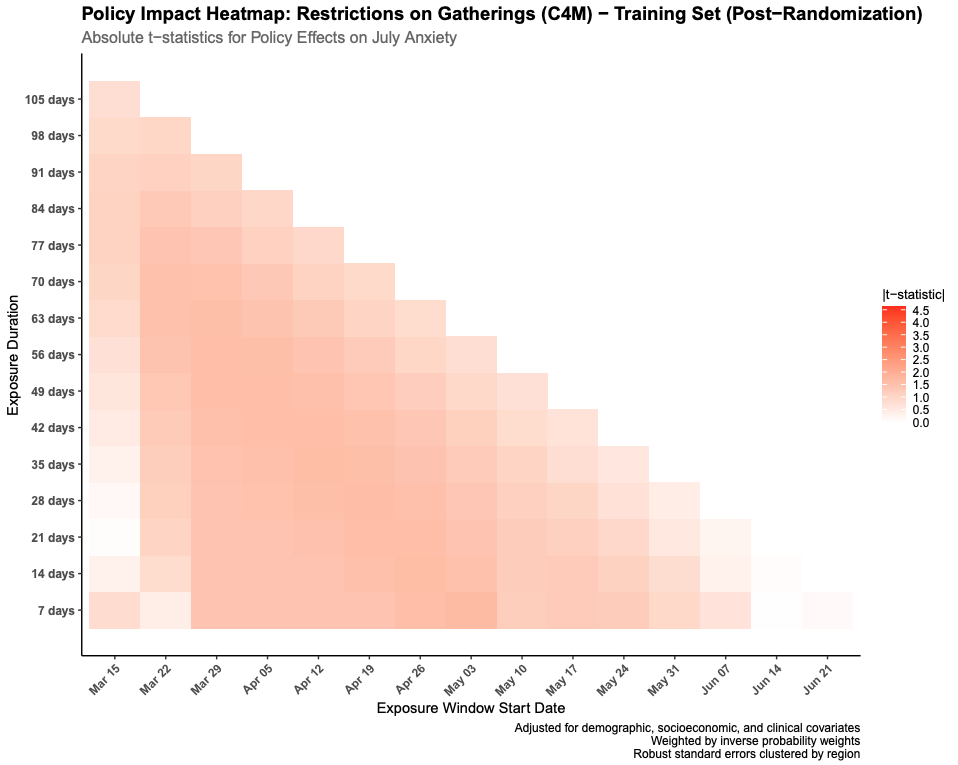

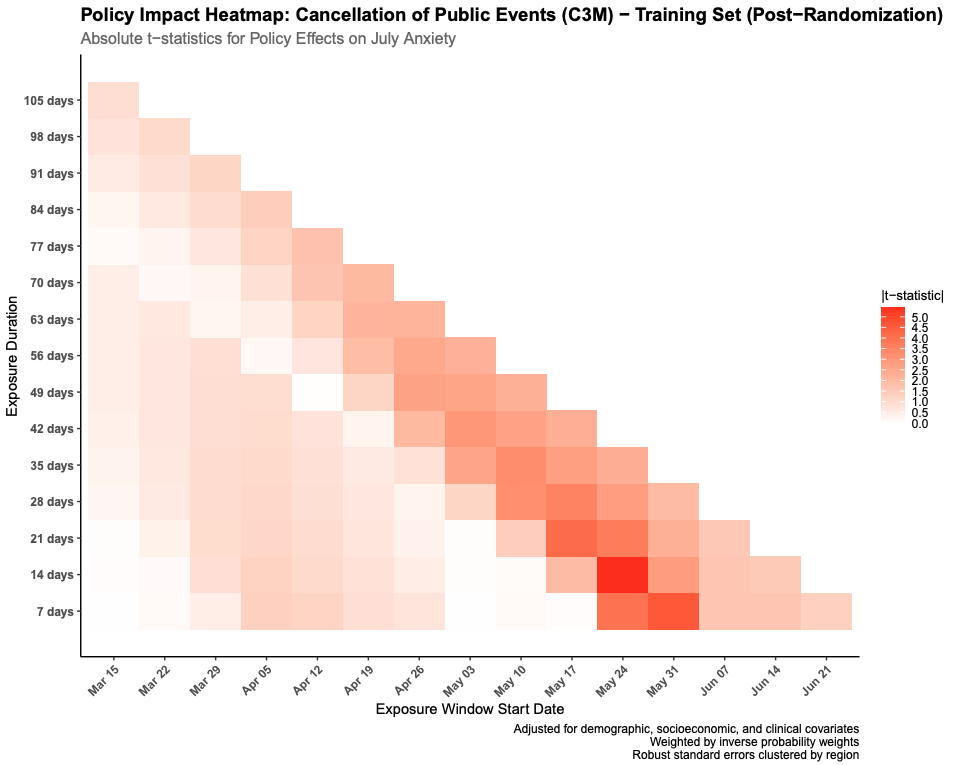

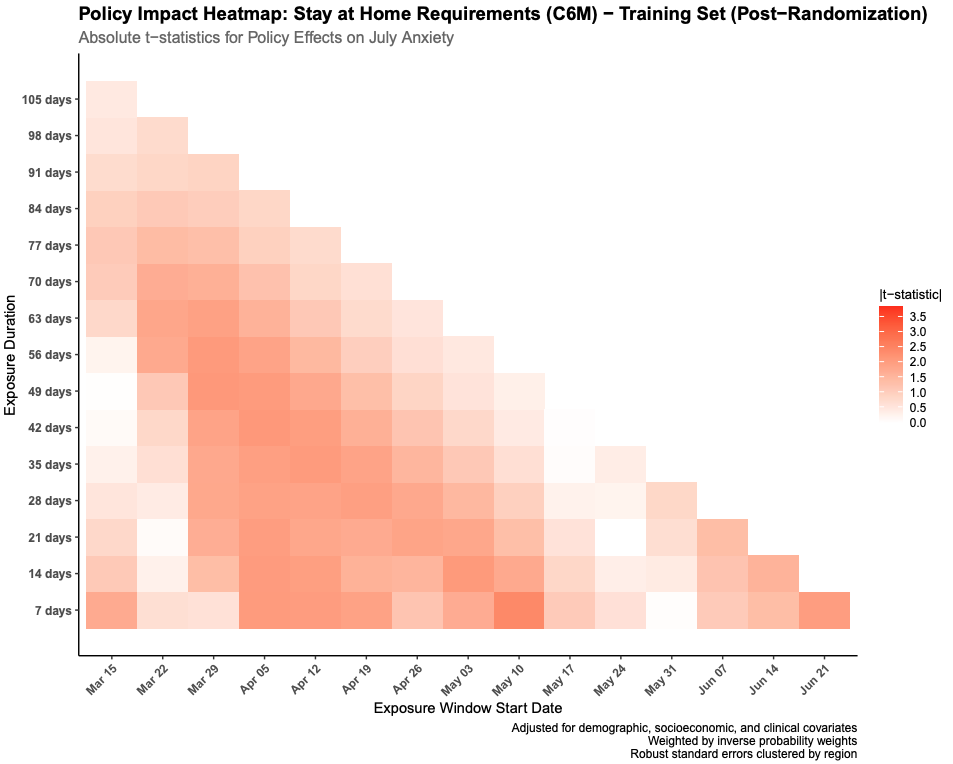

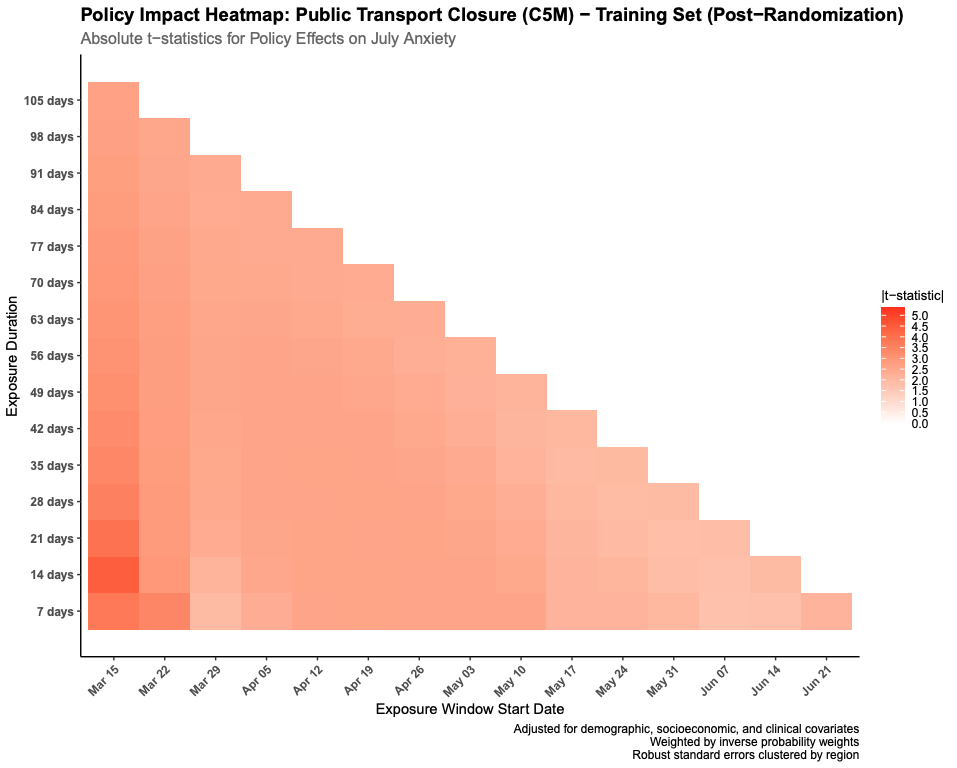

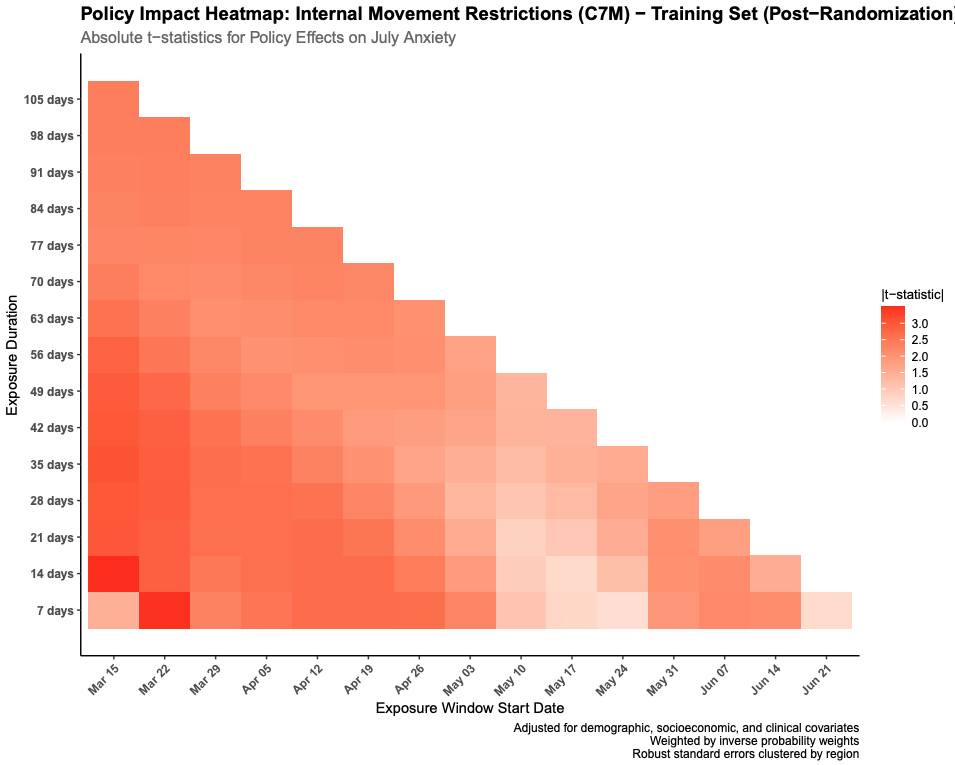

**eFigure 7. Policy-specific t-statistic heatmaps revealing temporal patterns of containment policy-anxiety associations across exposure windows.** Individual absolute t-statistic heatmaps for each of the seven COVID-19 containment policies (C1M-C7M) across 120 potential exposure windows, derived from linear regression models using training data (n=20,288) to quantify associations between 7-day policy exposures beginning from March 15th through June 21st, 2020 and self-reported GAD-7 anxiety symptoms in July 2020. Each heatmap displays t-statistics on its policy-specific scale to prevent scale compression that would obscure temporal patterns in weaker-associating policies when applying a uniform scale across all seven containment measures. Cluster-robust variance estimation was used to account for potential correlation within states, and regression estimates were adjusted for demographic, socioeconomic, and history of pre-pandemic anxiety disorders.

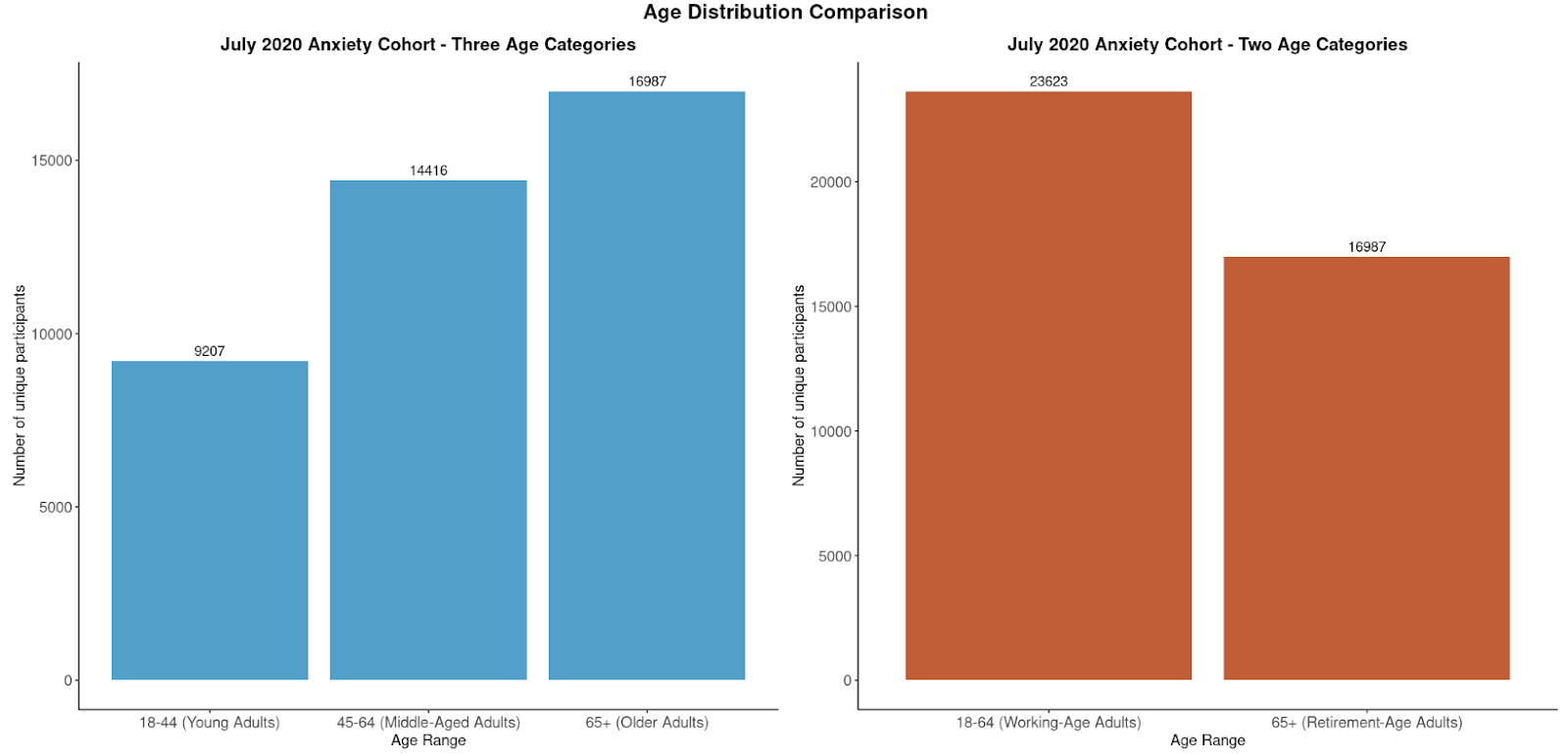

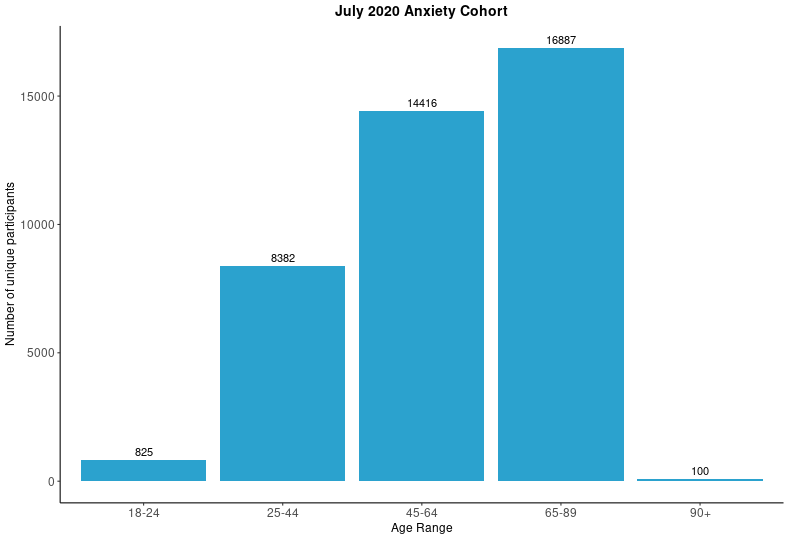

**eFigure 8. Age distribution of study participants and age stratification for subgroup analysis.** Distribution of participants by age in the *All of Us* Research Program who completed the GAD-7 July 2020 outcome (n=40,610) shown across detailed age ranges (Panel A) and collapsed into three age categories used for stratified analyses (Panel B). The three age categories used in age-stratified exposure mixture model analyses were defined as young adults (18-44 years, n = 9207), middle-age adults (45-64 years, n = 14,416), and older adults (65+, n = 16,987). These groupings were selected based on distinct social and economic vulnerabilities that could influence responses to containment policies. Younger adults (18-44) were hypothesized to show stronger associations due to greater reliance on workplace environments for social connection, and higher vulnerability to employment disruption. Middle-aged adults (45-64) were expected to experience differential impacts through caregiving responsibilities potentially complicated by school closures and social gathering restrictions. Older adults (65+) were anticipated to have distinct response patterns due to differences in social networks, employment status, and health risk considerations.

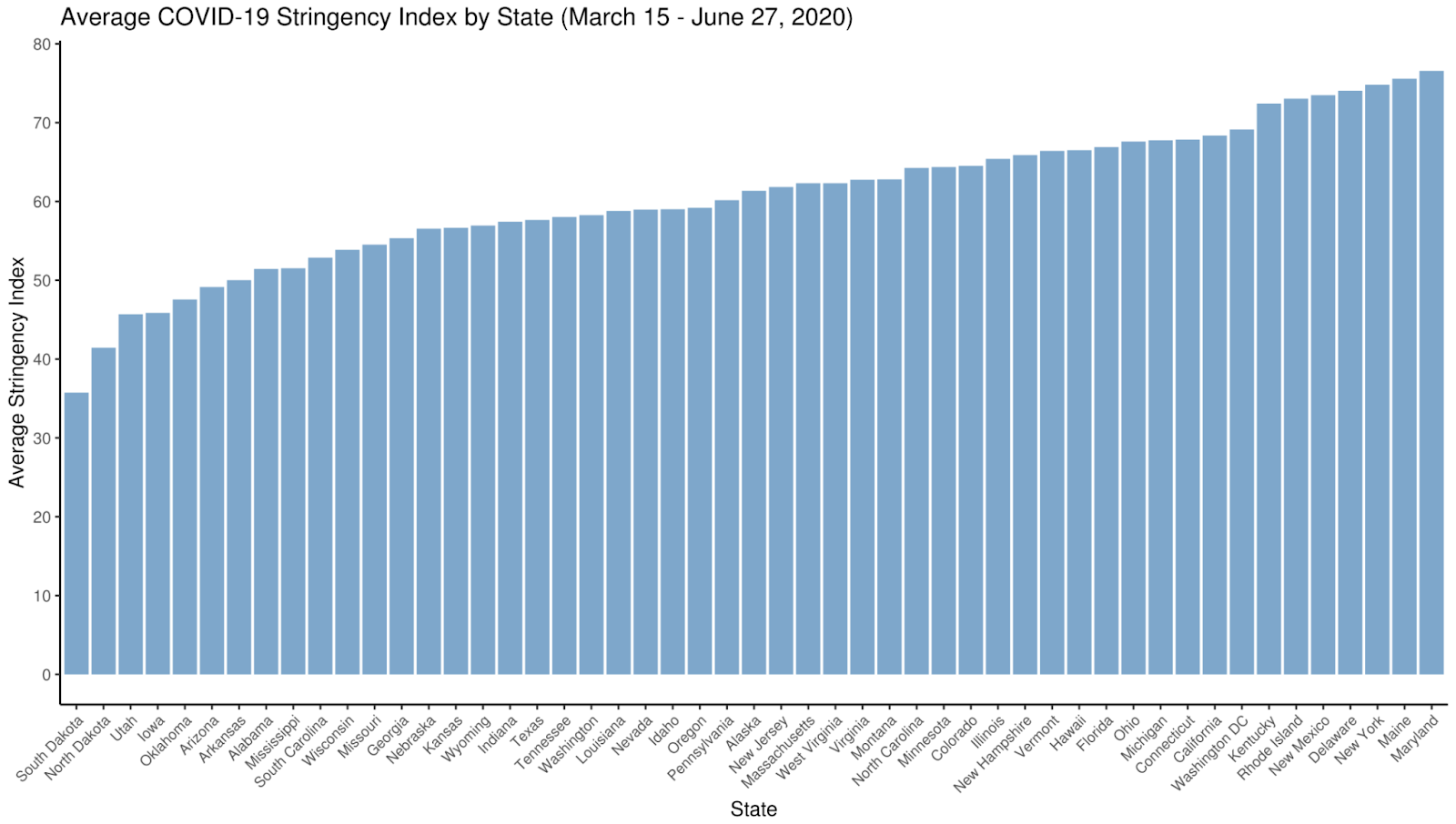

**eFigure 9. State-level variation in COVID-19 stringency index during the March 15th-June 27th, 2020 exposure window.** Average Oxford Coronavirus Government Response Tracker (OxCGRT) stringency index scores across all 50 US states and the District of Columbia during the March 15th - June 27th, 2020 exposure period. The stringency index is a composite measure (0-100 scale) of nine containment policy responses including school closures, workplace closures, cancellation of public events, restrictions on public gatherings, public transport closures, stay-at-home requirements, public information campaigns, internal movement restrictions, and international travel controls. States with the lowest average stringency included South Dakota (35.73), North Dakota (41.44), Utah (45.69), Iowa (45.87), and Oklahoma (47.56), while states with the highest stringency included Maryland (76.57), Maine (75.58), New York (74.80), Delaware (74.06), and New Mexico (73.49). This variation in state-level policy stringency provided the exposure heterogeneity necessary for quantile g-computation exposure mixture models examining differential policy effects on anxiety outcomes.

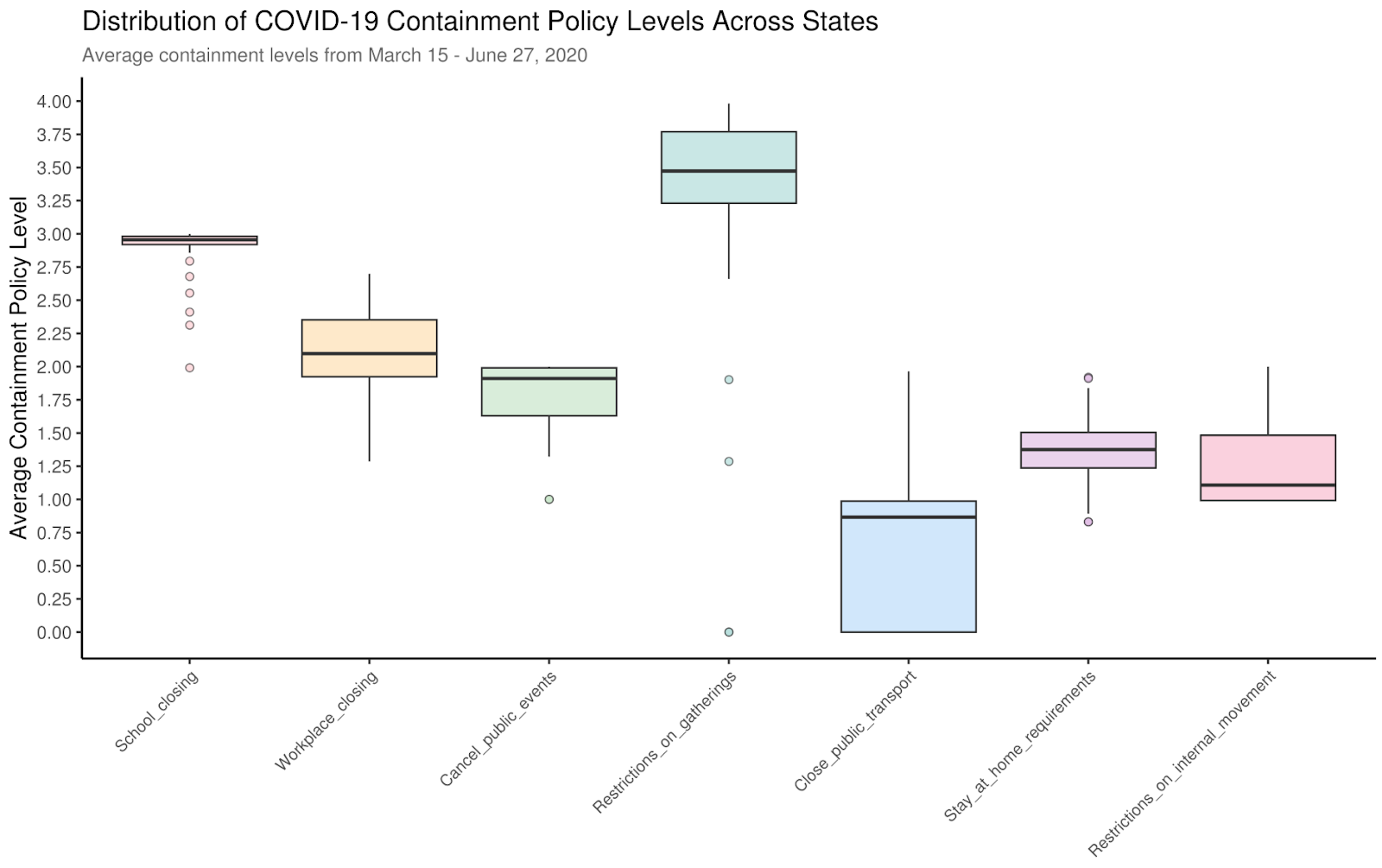

**eFigure 10. Distribution of average COVID-19 containment policy levels across US states during the March 15th - June 27th, 2020, exposure window.** Boxplots displaying the distribution of average containment policy implementation levels across all 51 states (50 states plus Washington, DC) for each of the seven COVID-19 containment policies during the March 15th-June 27th, 2020, exposure period. Policy levels represent continuous averages of ordinal implementation scales over the entire exposure window, capturing the intensity of policy implementation rather than discrete categorization at specific time points. Restrictions on gatherings demonstrated the highest average implementation levels across states (mean = 3.29) with substantial interstate variation (SD = 0.838, range: 0-3.98). School closures showed the most uniform implementation across states (mean = 2.90, SD = 0.188) with nearly all states maintaining high closure levels (range: 1.99-3.00). Public transportation closures exhibited the lowest average containment policy levels (mean = 0.719) and highest relative variation across states (SD = 0.59, range: 0-1.96), indicating substantial heterogeneity in this policy's adoption. Workplace closures (mean = 2.11), stay-at-home requirements (mean = 1.35), cancellation of public events (mean = 1.79), and internal movement restrictions (mean = 1.28) demonstrated intermediate containment policy levels with varying degrees of interstate heterogeneity. This variation in policy implementation across states and containment policies provided the necessary exposure heterogeneity for quantile g-computation mixture analyses examining differential effects on anxiety outcomes.

### eMethods

**Missing Data**

Missing covariate data were addressed using random forest imputation. Five imputation runs were performed using predictive mean matching with five nearest neighbors, each configured with 1000 trees and five iterations. Final imputed values were determined by consensus aggregation (mode for categorical variables, median for continuous variables), an approach that accommodates both data types without strict parametric assumptions while partially mitigating single-imputation bias.

**Containment Policies**

We excluded international travel restrictions because this measure showed minimal variation across states during our study period (March 15^th^ – July 31^st^, 2020). Each containment policy was measured on an ordinal scale capturing increasing levels of policy restrictiveness. For example, gathering restrictions ranged from 0 (no restrictions) to 4 (restrictions on gatherings of 10 people or less), while stay-at-home requirements ranged from 0 (no measures) to 3 (requiring staying home with minimal exceptions). School and workplace closures similarly ranged from 0 (no measures) to 3 (requiring closing for all levels or all non-essential workplaces), with lower values capturing recommendations or partial requirements. OxCGRT codes the most restrictive policy in effect within each state, capturing jurisdiction-wide implementation rather than policies limited to specific localities. We also examined the OxCGRT stringency index, a prespecified sum of nine containment indicators (0-100 scale), rescaled to 0-4 to match individual policy ranges.

**Identifying exposure windows with the highest containment policy-anxiety associations**

To identify exposure windows during which policy-anxiety associations were strongest, we conducted sequential evaluation of 120 potential windows defined by 7-day incremental start dates from March 15th through June 21st and varying durations across the 105-day candidate exposure period (March 15 - June 28, 2020). To prevent overfitting during window selection, we split the analytic sample into training (n=20,288) and testing (n=20,322) sets using rerandomization. We evaluated 100 candidate partitions and selected the split with the lowest area under the curve (AUC) from logistic regression models predicting set membership based on baseline covariates, ensuring maximal covariate balance between sets. AUC ranged from 0.511-0.527, resulting in a 1.8% improvement in balance relative to using a simple stratified training and testing split. Each rerandomization used stratified sampling to maintain balanced distributions across geographic regions (with 15 low-prevalence states collapsed into three groups based on state-level stringency tertiles; eFigure 1) and baseline anxiety severity (categorized into tertiles: none [0–1, n=10,841], minimal [1–5, n=14,006], and mild/severe [5–21, n=15,763]; eFigure 2). Balance was assessed using chi-square tests for geographic regions (eFigure 1), Kolmogorov-Smirnov tests for policy distributions (eFigure 2), standardized mean differences for both continuous and binary covariates (eFigure 3), as well as policy exposures (eFigure 4), and Cramer’s V for categorical variables (eFigure 5).

For each window, we fitted linear regression models with cluster-robust standard errors, adjusting for demographic, socioeconomic, and pre-pandemic anxiety covariates. This systematic evaluation revealed substantial heterogeneity in both temporal patterns and strength of individual policy associations with anxiety. Based on this evaluation, we selected two exposure windows for quantile g-computation analyses. The full exposure window (March 15-June 28th, 2020; 105 days) captures the complete period from initial policy implementation through early summer. The primary data-adaptive window (March 22-May 23, 2020; 63 days) was selected because policy-anxiety associations were strongest during this period, which coincides with the acute implementation phase when policies were most stringent and population exposures to restrictions were higher across states. This window excludes the initial week when policies were just being enacted and the later period when restrictions began easing and populations may have adapted to sustained measures. As an additional sensitivity analysis, we examined a shorter data-adaptive window (April 12-May 23, 2020; 42 days) to assess whether mixture effects were robust when restricting to the period of peak policy stringency (eTable 7). This window excludes the transition period in late March and early April when policies were still being implemented across states, isolating the interval during which containment measures were most consistently enforced nationwide. Comparison of effect estimates across these windows tests whether the observed associations depend on the inclusion of the early implementation period or whether they persist when restricting to the most stringent phase.

**Cluster-robust standard errors**

All analyses used cluster-robust standard errors to account for within-state correlation arising from state-level policy exposures and individual-level anxiety outcomes across 50 states.²⁰ We implemented the CR3 small-sample adjustment via the clubSandwich package in R, which provides bias-reduced linearization estimators appropriate when the number of clusters is moderate.²¹ The CR3 adjustment applies a correction that improves coverage of confidence intervals when cluster sizes are unequal, as occurred in our data where state-level sample sizes ranged from 85 to 6233 participants. Standard errors were clustered at the state level because containment policies were implemented at this level, inducing correlation among participants within the same state who experienced identical policy exposures.

**COPE non-response inverse probability weighting**

Missing covariate data were addressed via random forest imputation. Probabilities were trimmed at a minimum of 0.001.

**Observed confounding**

For exposure mixture models, we adjusted for a targeted set of potential confounders. Demographic variables included age, sex, race/ethnicity, birthplace, and marital status. Socioeconomic indicators encompassed education, employment status, and health insurance status. We incorporated pre-pandemic anxiety measures using Systematized Nomenclature of Medicine Clinical Terms (SNOMED)^15^ diagnostic codes from participants’ EHRs, including any anxiety disorder (48694002) or generalized anxiety disorder (21897009). Pre-pandemic anxiety was coded as two binary indicators, one for lifetime history prior to March 15^th^ 2020, and another for past-year diagnoses (March 15^th^ 2019 - March 15^th^ 2020). These covariates were selected based on their potential to confound the relationship between containment policies and anxiety outcomes. For the inverse probability weighting model, we incorporated both the covariates from our exposure mixture models and additional pre-pandemic characteristics from the *All of Us* Basics Survey including country of birth, race/ethnicity, age at enrollment, sex assigned at birth, income, education, primary language, and sexual orientation. For categorical variables with rare values, such as country of birth, we collapsed categories with fewer than 100 observations to ensure model stability. We also included relevant health indicators based on SNOMED codes from participants’ EHR data. Each health condition was coded as two binary indicators, one for past-year presence (prior year before March 15^th^, 2020) and another for lifetime history prior to March 15^th^, 2020. These conditions included hypertension, inflammatory neuropathy, congestive heart failure, mood disorders, anxiety disorders, sleep disorders, obesity, diabetes, high cholesterol, asthma, breathing difficulties, bronchitis, respiratory tract disorders, and autoimmune diseases.
